# Intraoperative copy number profiling from ultra-low coverage long-read sequencing for molecular tumor assessment

**DOI:** 10.64898/2026.07.21.26358307

**Authors:** Gaojianyong Wang, Caroline Kubelt, Romualdas Smičius, Kevin Zidane, Christian Rohrandt, Björn Brändl, Derek Wong, Mara Steiger, Amy Lum, Maxmilian Evers, Nils O. Schmidt, Martin Pröscholdt, Markus J Riemenschneider, Helene Kretzmer, Michael Synowitz, Stephen Yip, Martin Vingron, Franz-Josef Müller

**Affiliations:** Department of Bioinformatics, Max Planck Institute for Molecular Genetics, Berlin, Germany; Department of Genome Regulation, Max Planck Institute for Molecular Genetics, Berlin, Germany; Department of Psychiatry and Psychotherapy, Christian-Albrechts University, Kiel, Germany; Institute for Communications Technologies and Embedded Systems, Kiel University of Applied Sciences, Kiel, Germany; Hasso Plattner Institute for Digital Engineering, Digital Engineering Faculty, University of Potsdam, Potsdam, Germany; Department of Neurosurgery, University Medical Center Schleswig-Holstein UKSH, Campus Kiel, Kiel, Germany; Molecular Oncology, BC Cancer, Vancouver, BC, Canada; Department of Pathology, Children’s Hospital of Philadelphia, PA, USA; Altona diagnostics GmbH, Hamburg, Germany; Institute for Biology and Biotechnology of Plants, University of Münster, Münster, Germany; Department of Neurosurgery, University Medical Center Regensburg, Regensburg, Germany; Brain Tumor Center, University Medical Center Regensburg, Regensburg, Germany; Department of Neuropathology, University Medical Center Regensburg, Regensburg, Germany; Department of Pathology & Laboratory Medicine, Faculty of Medicine, University of British Columbia

**Author notes:** Equal contribution (Martin Vingron, Franz-Josef Müller).

**Keywords:** copy number variation, glioma, intraoperative diagnosis, cancer stratification

## Abstract

Copy number variations (CNVs) can serve as important clinical biomarkers for tumor classification and stratification. However, the utility of these CNV biomarkers for intraoperative tumor assessment within the timeframe of neurosurgical procedures has remained elusive due to the protracted duration of conventional CNV characterization methods. Here, we introduce CNVisor, a statistical framework for reliable and robust CNV detection from long-read sequencing, even under ultra-low coverage. Applied to neurosurgical tumor specimens, the proposed method enabled genome-wide CNV profiling and identified clinically relevant CNVs using roughly 60,000 reads within 20 minutes of sequencing. Integrating CNVisor with methylation-based classifiers can further reduce turnaround time and increase the accuracy of glioma subtype stratification. Together, these findings establish real-time CNV profiling using ultra-low coverage nanopore sequencing as a feasible strategy for intraoperative, genomics-informed assessment of CNS tumors.

## Introduction

Cancer acquires the capability to divide uncontrollably through the gradual accumulation of somatic alterations [1, 2], including copy number variations (CNVs), where the copies of DNA regions change compared with a normal genome [3, 4]. Technologies used to identify CNVs have evolved over time, increasing in resolution from chromosomal-level karyotyping to kilobase pair (kbps) level next-generation sequencing (NGS) [5–9]. By using these approaches, recurrent CNVs among different cancer types have been found to be important biomarkers [10–15], guiding critical decisions in cancer diagnosis and treatment as well as monitoring treatment response. As a result, CNV characterization has become standardized in clinical sequencing workflows, patient care protocols and in clinical tumor boards’ decisions [16].

CNV characterization is particularly relevant in central nervous system (CNS) tumors, for which the 2021 WHO classification increasingly integrates molecular profiles with histopathology to define tumor entities and support diagnosis, prognosis, and risk assessment [17, 18]. Within adult gliomas, IDH-mutated oligodendroglioma can be differentiated from IDH-mutated astrocytoma by co-deletion of chromosomes 1p and 19q, which is associated with an improved response to chemotherapy and longer overall survival [19, 20]. Conversely, chromosome 7 amplifications (7+) and chromosome 10 deletions (10-) are associated with IDH-wildtype glioblastoma and a poorer prognosis [21, 22]. Additionally, gene-level CNVs are also important for glioma diagnosis and prognosis. EGFR amplification is associated with glioblastoma [21, 22]. CDKN2A/B homozygous deletion distinguishes IDH-mutant astrocytoma, CNS WHO grade 4, from lower-grade astrocytoma. Grade 2 or 3 IDH-mutated oligodendroglioma with CDKN2A/B homozygous deletion is linked to a particularly poor prognosis [23].

Since molecular glioma classification is often not available before surgery, intraoperative knowledge of glioma subtype and grade can be clinically beneficial for patients, as it informs clinical decisions regarding treatment planning and the extent of resection. For instance, extensive resection is more viable for gliomas with a good prognosis, e.g., IDH-mutated oligodendroglioma with 1p19q co-deletion but without CDKN2A/B deletion. Conversely, limited resection aimed at minimizing neurological damage could enhance the remaining quality of life of patients with an extremely poor prognosis, such as IDH-wildtype glioblastoma. Nanopore sequencing has enabled the interpretation and analysis of nucleotides and methylation status of DNA and RNA molecules in near real-time [24–29]. With this technology, diagnostic and prognostic cancer biomarkers can be detected intraoperatively using suitable live bioinformatic methods [25–30]. Recent nanopore-based methylation classifiers [26, 27] have demonstrated the feasibility of real-time epigenomic profiling for intraoperative tumor classification. Sturgeon [26] uses a neural network to classify CNS tumors from ultra-low coverage nanopore methylation data, whereas MethyLYZR [27] uses a naïve Bayesian framework for rapid, computationally tractable live classification.

However, glioma diagnosis and stratification require molecular information beyond methylation profiles, particularly the status of clinically relevant CNV biomarkers. Detecting these CNV biomarkers within an intraoperative timeframe remains technically challenging. Neuro-oncological procedures have a median duration of 179 minutes, ranging from 123 to 250 minutes [31]. The timeframe in which a neurosurgeon, following the assessment of a CNS tumor via craniotomy, can employ diagnostic information derived from a biopsy to guide the subsequent resection, is generally considered to be limited to less than one hour. Conventional CNV analysis methods, which take days to weeks, are not suited for intraoperative use where rapid diagnostic decisions are critical.

Recent studies have attempted to improve the utility of real-time CNV characterization using nanopore sequencing. Rapid-CNS2 [29] and Nano-GLADIATOR [30] focus on chromosomal-level CNVs, leaving gene-level CNVs, e.g., CDKN2A/B deletions or EGFR amplifications, undetected. Mechanically fragmenting DNA to infer CNVs at higher resolution was employed previously at the cost of quicker nanopore depletion [32]. SMURF-seq, ligates enzymatically fragmented DNA into concatenated long-reads to increase throughput and yield [33], and generates clinically actionable results within 4 hours [34]. iSCORED [35] improves upon SMURF-seq by employing the one-reaction enzymatic digestion approach and decreases the turnaround time down to 120-140 mins. However, these methods still do not fully align with the timeframe of neuro-oncological procedures, in which molecular information generally needs to be available within 60 minutes to guide subsequent resection. Expansion of this tight window to incorporate the above CNV analysis workflows is impractical, due to not only the risk of poorer patient outcomes from extended surgeries [31] but also resource limitations in even the most well-equipped tertiary centers.

Beyond turnaround time, the integration of CNV biomarkers into intraoperative tumor classification has been, so far, unfeasible due to the lack of sensitivity and reliability of CNV detection in smaller genomic regions, such as 19q [25, 26, 36], let alone the gene-level CNVs. This limitation arises because conventional CNV detection algorithms are primarily tailored for NGS and optimized to identify CNVs at kbps resolution. For example, circular binary segmentation (CBS) was designed to address fluctuation of CNVs in NGS by combining adjacent genomic bins with similar signals into larger segments [37]. It performs well with kbps-level bins. Using the similar bin settings with long-read sequencing often demands a coverage that exceeds NGS by more than two orders of magnitude, which is impractical in any clinical setting. This segmentation algorithm then becomes problematic when the bin size is increased to the Mbps level, since it can obscure gene-level CNVs through bin settings or by merging adjacent bins without CNVs into the same segment.

Importantly, a pan-cancer study of over 10,000 samples has revealed that 97% of all detected CNVs are larger than 100 kbps (**Fig. S1**) [15], suggesting that intraoperative genomic analysis targeting clinically relevant CNVs does not always require the higher resolution provided by NGS. Clinically relevant CNVs may be detectable from ultra-low coverage sequencing data when the statistical framework explicitly models uncertainty in the observed CNVs and resolves CNV status once sufficient evidence has constrained this uncertainty to an acceptable level, without requiring the exact CNV values.

In this study, we address limitations of the existing methods and offer a real-time CNV characterization solution suitable for clinical settings with the potential to improve diagnosis and treatment decisions. With CNVisor, we describe a statistical framework to determine the minimum sequencing time required for genome-wide CNV characterization, to narrow down CNV breakpoints, and to provide a robust CNV range assessment. We validate our method on human cell lines and glioma biopsies processed in our laboratory and applied it to eight brain tumor patients during surgery. Our findings demonstrate CNVisor’s capability to reliably detect CNVs within 20 minutes of sequencing (60 minutes after receiving a biopsy). Furthermore, we demonstrate that integrating CNVisor with a methylation-based glioma classifier can significantly enhance intraoperative diagnostics by reducing the required sequencing time and increasing the accuracy of glioma subtype stratification.

## Results

### Statistical framework for live CNV characterization

Nanopore sequencing can be conceptualized as a stochastic sampling of reads from the genome with the number of reads originating from a genomic region following a binomial distribution **(Fig. 1a**, **Materials and Methods, Supplementary Document**). Given that the sequencing coverage achievable within the clinically relevant timeframe of less than one hour from biopsy [25–27] is low compared to that from a full sequencing run, which typically extends to 72 hours [38, 39], we make two assumptions. First, the probability of obtaining reads from a genomic region is not affected by the number of reads already obtained from the same region. Second, obtaining of reads from different regions is independent of each other.

**Fig. 1.**
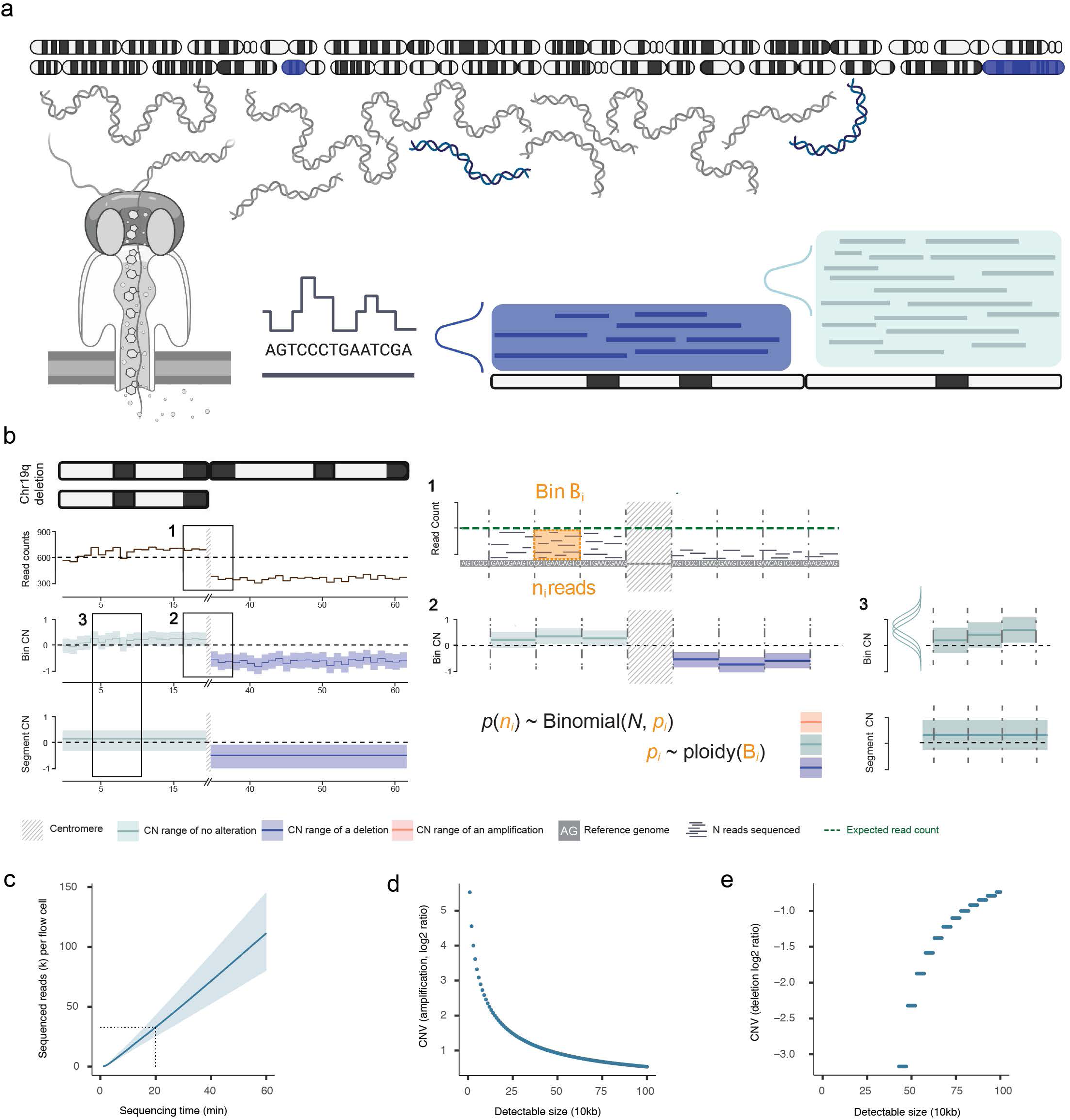
Live CNV characterization. (a) Illustration of live CNV characterization. Nanopore reads from a genome with 1p/19q co-deletion are basecalled and aligned to the reference genome to obtain the read distribution. (b) CNVisor analysis exemplified from an oligodendroglioma biopsy (IEG4). Read counts from sparse sequencing data are plotted. 1) Counting reads in each bin. 2) CN and its range of each bin are computed by CNVisor. 3) Based on likelihood functions, neighboring bins with similar CNs are merged into the same segment. The range of the segment is adjusted by Bonferroni correction. (c) Relationship between the number of sequenced reads per flowcell and sequencing time. The blue line represents the average number of sequenced reads of 20 flowcells and the blue background is the range of sequenced reads of all 20 flowcells. (d)-(e) Simulation of the detectable CNVs, assuming that 60,000 reads are sequenced (average sequenced reads of two MinION flowcells in 20 minutes) and the genome is evenly split into bins of 3 Mbps. Assuming that a bin adjacent to a diploidy bin contains CNVs smaller than the bin size. The relationship between CNV sizes and the minimum amplification that can be detected (d). The relationship between CNV sizes and the maximum deletion that can be detected (d).

The characterization of a genomic region for CNVs can then be understood as the disparity assessment between the observed and expected numbers of reads from this region (**Fig. 1 b-1**). The reference genome is partitioned into non-overlapping bins (**Fig. S2, Materials and Methods, Supplementary Document**), and the number of reads aligned to each bin is compared with its expected value to determine its copy number (CN). In ultra-low coverage nanopore sequencing, CN of each bin is usually unstable. We model CN as ploidy factor (**Materials and Methods, Supplementary Document**). Within each bin, the likelihood of different ploidy factors, given the observed number of aligned reads in this bin and the total number of sequenced reads, can be written in the form of a binomial distribution (**Materials and Methods, Supplementary Document**). This likelihood function follows a beta distribution after normalization and we can compute the range for ploidy factors accordingly (**Fig. 1b-2, Materials and Methods, Supplementary Document**).

**Fig. 2.**
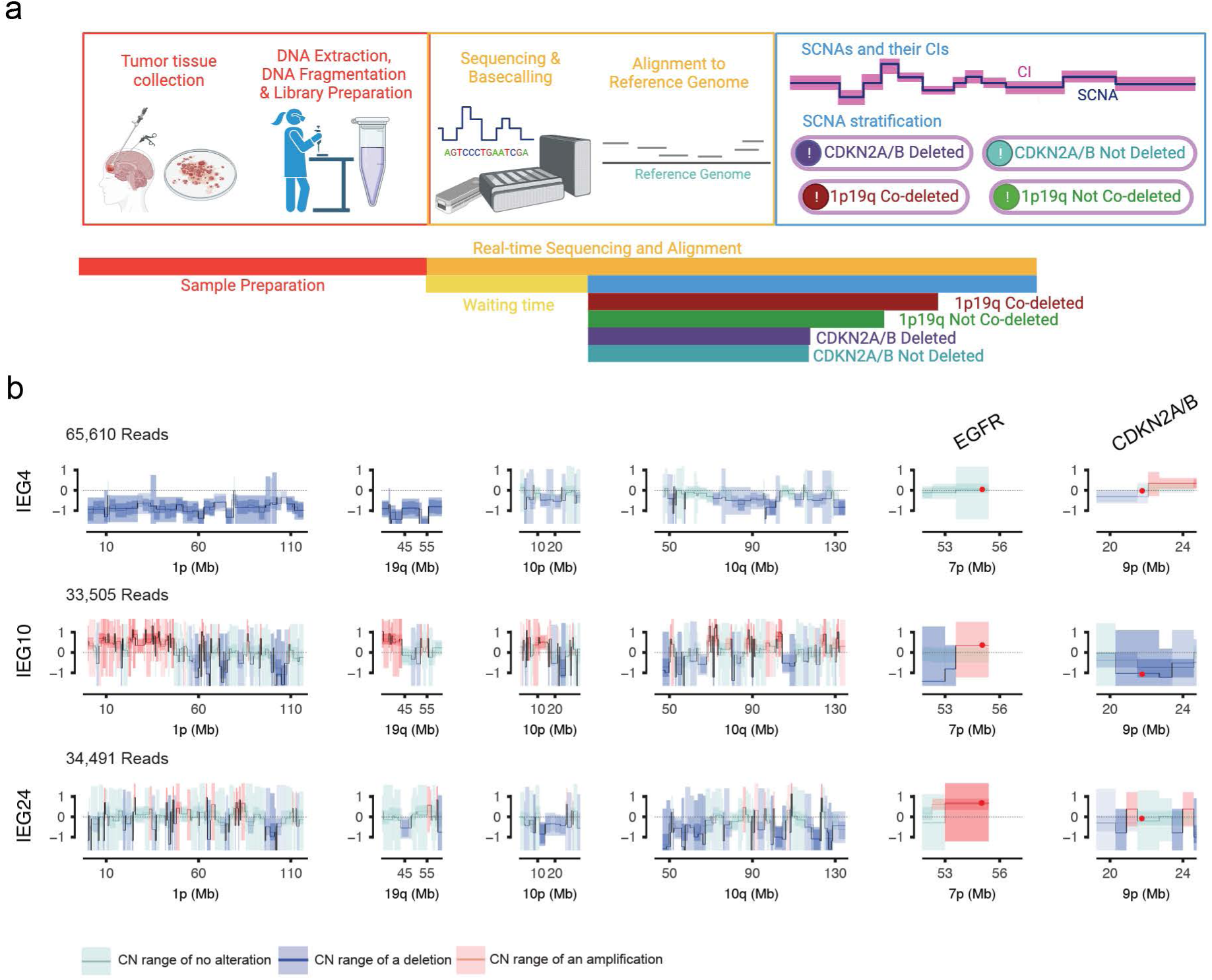
Intraoperative CNV characterization procedure. (a) Integrated surgical, molecular, and bioinformatic process for CNV assessment at the Point-of-Care (b) CNV of three glioma samples, IEG4 (oligodendroglioma), IEG13 (astrocytoma), and IEG24 (glioblastoma) at 1p19q, chr10, EGFR and CDKN2A/B. X-axis is the regions of interests and y-axis is log2 ratio CN. Blue, red, and light green respectively represent amplification, deletion, and no alteration. The colored line is the CNV of the bin/segment, and the colored shade is its CNV range.

We then used the joint likelihood function between neighboring bins to evaluate whether the ploidy factors of neighboring bins are statistically similar. A breakpoint is introduced between two bins if their ploidy factors are statistically different (**Materials and Methods, Supplementary Document**). The bins between two breakpoints that exhibit similar CNs are merged into larger segments. Since the breakpoints can be biased by bin settings, we optimize CNV breakpoints through a dynamic strategy. The bins on both sides of each initial breakpoint are further divided into sub-bins to identify the optimal breakpoints (**Materials and Methods, Supplementary Document**). After breakpoint optimization, we adjust the ranges of these segments using Bonferroni correction (**Fig. 1b-3, Materials and Methods, Supplementary Document**).

**Fig. 3.**
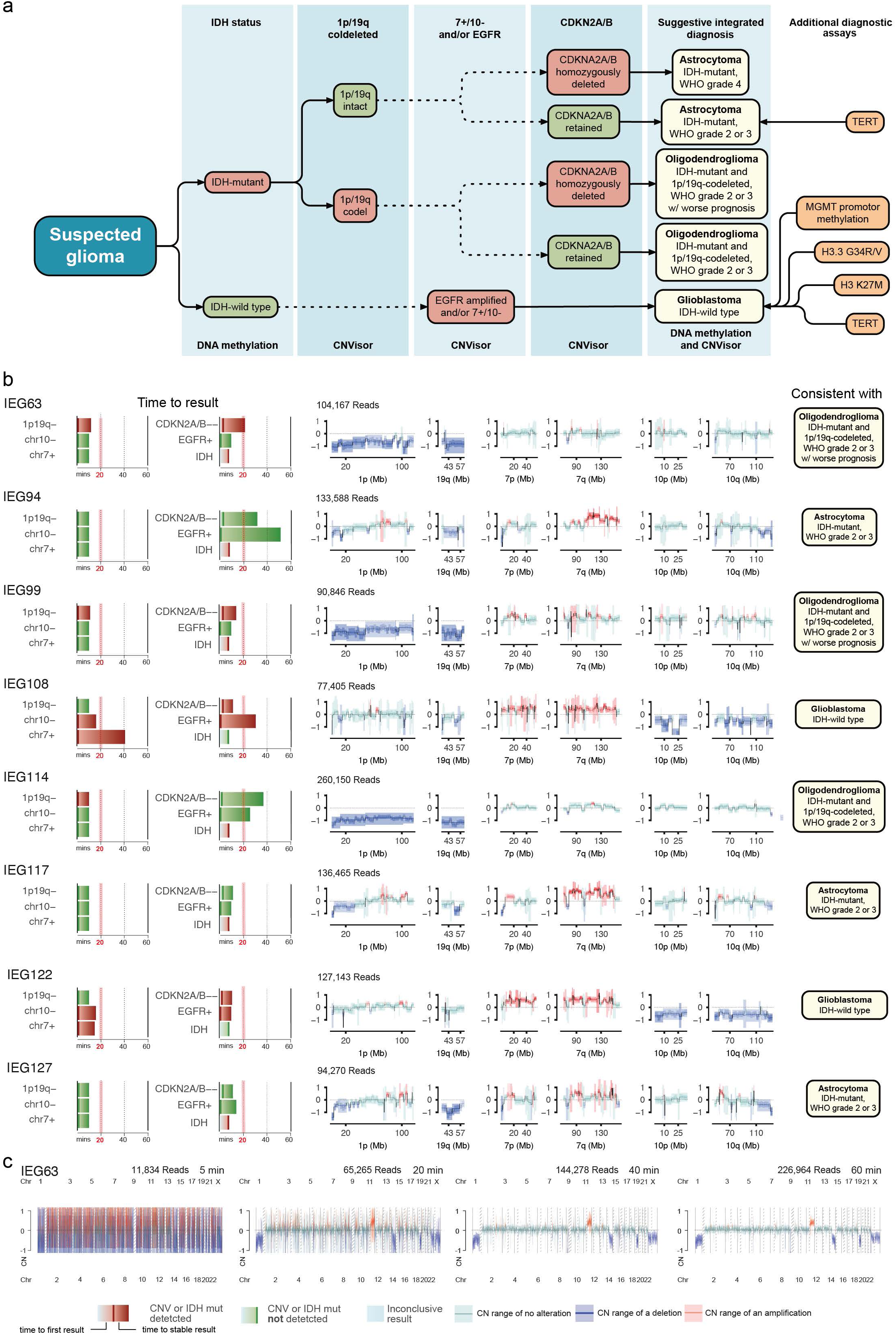
Intraoperative utilization of CNVisor facilitating glioma diagnosis. (a) The differentiation between major diffuse gliomas in adults with CNVisor adapted from the EANO diagnosis and treatment guidelines of diffuse gliomas in adults [18]. (b) CNVisor results of eight surgical specimens and corresponding suggested diagnosis within 1 hour of sequencing. Results are plotted in pseudo-time after a flow cell has been loaded with a library. The time required to obtain the CNV status of regions of interests (1p19q, co-deletion, 7+, 10-, CDKN2A/B homozygous deletion, and EGFR amplification) are illustrated on the left. The CNVs of these regions are illustrated in the middle. The suggested diagnosis from CNViosr is illustrated on the right. (c) CNVisor results from a time series (5, 20, 40, and 60 minutes) of the sample IEG63.

In addition to the CNV ranges, we also obtain the likelihoods of alterations for all resulting segments to provide an automatic quantification of CNVs **(Supplementary Document**). The level of amplifications and deletions, e.g., heterozygous deletions of 1p19q or homozygous deletions of CDKN2A/B, can be estimated from the obtained likelihoods.

Next, we conducted statistical analyses and demonstrated the feasibility of CNV characterization at genomic regions of diagnostic relevance within one hour after obtaining an intraoperative biopsy (**Fig. S3, Supplementary Document**). A MinION sequencer requires only one minute of sequencing to generate sufficient reads for starting CNV characterization using CNVisor, with at least one aligned read in most of the bins. (**Fig. S3a, Fig. S3b, Supplementary Document**). With as few as 60,000 reads that can be obtained in an intraoperative setting with two MinION flow cells in parallel within 20 minutes of sequencing, our simulations have demonstrated that CNVisor can detect heterozygous deletions of 770 kbps, homozygous deletions of 450 kbps, and tetra-amplifications of 150 kbps (**Fig. 1c-e, Supplementary Document**). Extending the sequencing time to 60 minutes with up to 150,000 sequenced reads detected heterozygous deletions of 270 kbps, homozygous deletions of 160 kbps and tetra-amplifications of 70 kbps (**Fig. S3**).

### Intraoperative CNV characterization procedure and validation

Toward a clinically relevant intraoperative CNV characterization, we envision an integrated surgical, molecular, and bioinformatics process (**Fig. 2a**). Following the extraction and preparation of a biopsy obtained at a glioma surgery, the tumor DNA is sequenced on nanopore devices in parallel with live bioinformatic analysis. After biopsy and library preparation, the CNV characterization is started when the first batch of reads has been obtained, usually within less than one minute (**Fig. 2a, Supplementary Document**).

We first evaluated the performance of CNVisor using publicly available nanopore data from peripheral blood [40] and two glioma cell lines with karyotypic abnormalities, LN229 and U251MG, sequenced in our laboratory (**Materials and Methods, Supplementary Table 1**). The CNV results of the Ashkenazi Trio data demonstrated a good agreement with the expected diploid genome status of all three samples (**Fig. S4**). The results relating to the sex-chromosome were also consistent with their known biological information, HG002 (**Fig. S4a-b**) and HG003 (**Fig. S4c-d**) being karyotypically male and HG004 female (**Fig. S4e-f**). CNVs detected in glioma cell lines (**Fig. S5**) have a good agreement between the two sequencing runs for each cell line. Within each cell line, we extended the method for assessing similarity between CNV profiles described in [41] to compare CNV profiles obtained from CNVisor and the Illumina EPIC array, which showed significant similarity (P<10^-4^, **Fig. S5, Materials and Methods, Supplementary Table 2**). Furthermore, the identified alterations were consistent with those reported in a recent study characterizing CNVs of these two cell lines (**Fig. S5, Supplementary Document**) [42].

Next, we evaluated CNVisor with glioma samples that were sequenced with PromethION flowcells with high coverage and a ligation sequencing protocol to simulate live analysis through subsampling of reads (**Materials and Methods, Supplementary Table 1**). The samples included an oligodendroglioma, IEG4, an anaplastic pleomorphic xanthoastrocytoma, IEG10, and a glioblastoma, IEG24.

We inspected the CNV profiles of these samples using nanopore data obtained within the first 15 minutes of sequencing (approximately 33,000 – 66,000 reads) since we found that the number of sequenced reads obtained in the first 15 minutes with one R9 PromethION flowcell is similar to that obtained with two R9 MinION flowcells in 20 minutes (**Fig. 1c**, **Fig. 2b**). The CNV profiles obtained from subsampled reads agreed with the matched CNV results derived from Illumina EPIC arrays (**Fig S6**). Significant similarity between the two was observed as early as three minutes from start of sequencing (P<10^-4^**, Materials and Methods, Supplementary Table 2**).

Using CNVisor, 1p19q co-deletion could be identified in IEG4 (**Fig. 2b, Fig. S6a**) within 15 minutes of sequencing from one PromethION flowcell. Conversely, the absence of the 1p19q co-deletion in IEG10 and IEG24 could also be confirmed (**Fig. 2b, Fig. S6b-c**). Additionally, the deletion of chromosome 10 in IEG24 was also detected with the same experimental setup (**Fig. 2b, Fig. S6b**).

### Intraoperative CNV characterization for glioma patients

CNVisor was applied to eight brain tumor biopsies, sequenced on-site at the neurosurgical unit at University Hospital Schleswig-Holstein Campus Kiel using at least two MinION sequencers in parallel (**Materials and Methods, Supplementary Table 1**). The surgical procedure was carefully performed, allowing for the sequencing of tumor tissues as pure as possible [27]. The sequencing process utilized nanopore sequencing’s timestamps on reads. This feature facilitated the reproduction and subsequent visualization of our method’s CNV detection and tumor type classification capabilities with progressing sequencing time **(Video S1).** We focused on these regions of interest: 1p19q, chromosomes 7 and 10, EGFR, and CDKN2A/B. Each region yields one of three outcomes: ‘CNV detected’, ‘CNV not detected’, or ‘inconclusive’. Once a result is obtained, the CNV’s presence or absence is verified over a period of ten minutes, during which the CNV status remains unchanged. For regions that remain inconclusive during surgery, the genome-wide CNV profile, the CNV status of other regions of interest, and any available molecular or clinical information should be interpreted together within the clinical workflow (see section “Enhanced glioma diagnostics through integration of CNV and methylation”).

To demonstrate an intraoperative workflow, we combined methylation classification [27] with CNVisor results, following recent guidelines [18] and a decision tree proposed by Wongsurawat et al. [34]. This decision tree utilizes global methylation patterns affected by IDH1/2 mutations to differentiate glioma subtypes [26, 27, 43]. IDH mutations alter DNA methylation, distinguishing glioblastoma lacking these mutations, from lower-grade gliomas such as IDH-mutated astrocytoma and IDH-mutated 1p19q co-deleted oligodendroglioma [17, 44]. Following the indirect assessment of IDH1/2 mutation status, CNVisor’s identification of specific CNVs aids in further distinguishing between glioblastoma, oligodendroglioma, and astrocytoma **(Fig. 3a)**. Additionally, gene-level CNV calling, such as for CDKN2A/B, helps distinguish between grade 2/3 and grade 4 astrocytomas, or identifies grade 2 or 3 oligodendrogliomas with CDKN2A/B homozygous deletions, linked to a particularly poor prognosis [23]. It is important note that IDH mutation status alone does not exclude all differential diagnoses for a suspected glioma, necessitating further downstream molecular analysis (e.g., pTERT mutation status, H3.3.G34R/V status, H3 K27M status) in certain cases.

CNVisor accurately identified CNVs in all eight samples, aligning with integrated diagnostic outcomes **(Fig. 3b, Fig. S7, Fig. S8, Supplementary Table 3**). CNV profiles generated by CNVisor were significantly concordant with EPIC array–derived profiles three minutes from start of sequencing (P<10^-4^**, Materials and Methods, Supplementary Table 2**). It pinpointed chromosomal abnormalities, such as 1p19q co-deletion in IEG63 **(Fig. 3b, Fig. S7a**), IEG99 **(Fig. 3b, Fig. S7c),** and IEG114 **(Fig. 3b, Fig. S8a)**, and chromosome 10 deletion in IEG108 **(Fig. S3b, Fig. S7d)** and IEG122 **(Fig. S3b, Fig. S8c),** alongside gene-level abnormalities including CDKN2A/B homozygous deletions in IEG63 (**Fig. S7a**), IEG99 (**Fig. S7c**), IEG108 (**Fig. S7d**), and IEG122 (**Fig. S8c**), and EGFR amplifications in IEG108 (**Fig. S7d**) and IEG122 (**Fig. S8c**).

The analysis determined the subtype and grade of each sample through identified CNV biomarkers. The subtype and grade obtained from CNVisor and MethyLYZR are in good agreement. Results of glioma subtype assignment stabilized within 20 minutes of sequencing for all samples **(Fig 3b, Supplementary Table)**. The time to detect the 1p19q co-deletion status was maximum 12 minutes. Similar detection times were observed for chromosome 7 amplification and chromosome 10 deletion, suggesting that CNVisor could resolve chromosomal CNVs from the first minutes of sequencing in most cases **(Fig 3b, Supplementary Table 3)**. In contrast, detection of gene-level CNVs is more challenging, requiring longer sequencing time. CDKN2A/B and EGFR CNV statuses were respectively detected within 21 minutes in six and five samples, whereas the remaining samples required 32 to 52 minutes **(Fig 3b, Supplementary Table 3)**. Concurrently, MethyLYZR, a methylation classification algorithm was utilized to infer IDH1/2 mutation status and ascertain class membership among predefined CNS tumor classes in our intraoperative sequencing samples [27].

### CNVisor improves gene-level CNV detection compared with CBS

Effective intraoperative CNV profiling requires not only reliable chromosome-level CNVs, but also accurate detection of clinically relevant gene-level alterations. In glioma, this is particularly important for events such as EGFR amplification, which supports glioblastoma classification [18], and CDKN2A/B homozygous deletion, which is a marker of extremely poor prognosis [23].

However, CNV segmentation algorithms designed for NGS, e.g. CBS [37]., can obscure gene-level CNVs when the bin size is increased to Mbps-level due to two reasons. First, since focal CNVs can be smaller than the bin size, the observed CN value represents an average over the entire bin rather than the focal alteration itself. Second, the bin containing the focal CNV may be merged with neighboring bins containing no CNVs, further masking the gene-level alterations. We applied CBS to all eight samples and found that it did not pinpoint any CDKN2A/B homozygous deletion **(Fig. S9)**.

In comparison, CNVisor introduces breakpoints when adjacent regions are statistically different and further optimizes the breakpoints within each bin. This approach enables the identification of gene-level CNVs and allows us to accurately detect CDKN2A/B status in real-time for these samples.

### Sensitivity and specificity of CNVisor

In order to quantify the detection accuracy of CNVisor, we first generated deeply sequenced data for IEG63 and IEG127, which yielded 3.2 and 5.2 million aligned reads, respectively, after extending sequencing on two MinION flow cells over 72 hours. Subsampling the completed datasets allowed for the assessment of sensitivity and specificity for 1p19q co-deletions and CDKN2A/B homozygous deletions. Initially, 60,000 reads were sampled from each biopsy, reflecting the expected output of two MinION flowcells in 20 minutes **(Fig. 1c)**, and this was repeated 1,000 times. In these evaluations, sensitivity and specificity for 1p19q co-deletion and CDKN2A/B homozygous deletion were both at 100%. When sampling only 50,000 reads, both metrics for 1p19q co-deletions remained at 100%, whereas those for CDKN2A/B homozygous deletions slightly decreased to 99.9%. With a further reduction of sampling reads to 20,000 reads, the sensitivity for 1p19q co-deletions dropped to 99.3%, while specificity remained 100%, and CDKN2A/B homozygous deletions’ sensitivity and specificity were 98.4% and 96.8%, respectively. Therefore, for the pure tumor tissues carefully resected during surgery, CNVisor was able to achieve a high sensitivity and specificity in these clinical samples.

### Enhanced glioma diagnostics through integration of CNV and methylation

Since CNVs and methylation capture distinct molecular features, we next evaluated whether combining these profiles could improve intraoperative tumor classification and stratification, particularly in cases in which either molecular profile alone provides insufficient evidence. We collected a total of 123 tumor samples (**Supplementary Table 4**) comprising glioblastomas (63 samples), astrocytomas (32 samples), and oligodendrogliomas (28 samples) from the recent MethyLYZR study [27]. These samples were sequenced using rapid, multiplexed, and barcoded library preparation on PromethION R10 flowcells (sequencing protocols available in [27]). We processed all the sequencing reads through CNVisor and cross-referenced the resulting CNV profiles reported in MethyLYZR study [27] to establish the CNV ground truth for regions of interest, including chromosomes 1p19q, chromosome 7, chromosome 10, EGFR, and CDKN2A/B. Furthermore, we manually curated all the samples, confirming their subtypes using CNV profiles (**Supplementary Table 4**) in conjunction with methylation profiles from MethyLYZR.

We subsampled reads ranging from 1,000 to 150,000 to simulate intraoperative sequencing procedures (**Materials and Methods**) for each glioma. CNVisor-generated profiles were combined with MethyLYZR results to determine the glioma subtype in each subsampling. Based on the decision tree **(Fig. 3a)**, we developed CNVisor+, an unsupervised glioma classifier that integrates orthogonal CNV and methylation profiles **(Fig. 4a).** CNVisor+ prioritizes high-confidence results from either CNVs or methylation in cases of discrepancy between them (**Materials and Methods**). This classifier categorizes samples as ‘Astrocytoma’, ‘Oligodendroglioma’, ‘Glioblastoma’, or ‘Inconclusive’.

**Fig. 4.**
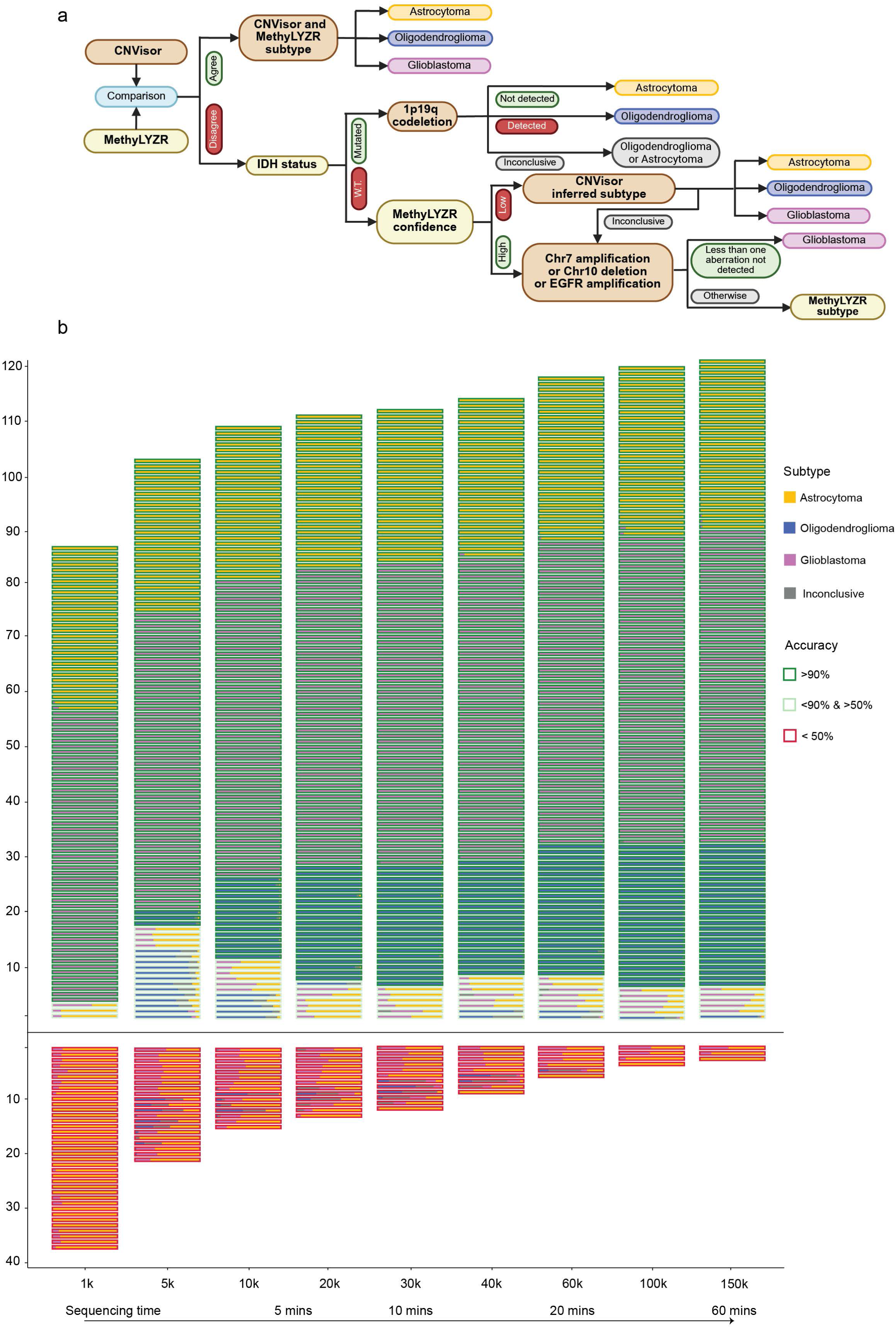
Enhanced glioma classification through integration of CNVisor and MethyLYZR. (a) The unsupervised classifier that integrates the CNV profiles from CNVisor and methylation profiles from MethyLYZR for accurate glioma diagnosis, which prioritizes high-confidence results from either CNV or methylation in cases of discrepancy between the two. It follows the diagnosis and treatment guidelines of diffuse gliomas in adults from the EANO [18]. (b) The accuracy of the unsupervised classifier using 123 glioma samples sequenced with multiplexed, barcoded PromethION R10 flowcells. To simulate the real-time sequencing process up to one hour, reads ranging from 1,000 to 150,000 were subsampled respectively for 100 times. The CNVisor-generated CNV profiles were paired with MethyLYZR results for glioma stratification. In the bar chart, the color of each bar represents the glioma subtype identified by the classifier, while the color of the rectangle surrounding the bars indicates the accuracy level.

We assessed the accuracy of CNVisor+ with different subsampled reads. With 60,000 reads, equivalent to 20 minutes of sequencing using two MinION flowcells, the classifier achieved >90% accuracy in 109 samples and >50% accuracy in 117 samples (**Fig. 4b**). Only 6 samples were misclassified, either incorrectly or marked as inconclusive, with an accuracy less than 50% (**Fig. 4b**). In contrast, using CNV profiles alone, we achieved >90% accuracy in 91 samples and >50% accuracy in 112 samples, with 11 samples mis-characterized. Methylation profiles alone only correctly classified 92 samples with high confidence and 5 samples with low confidence, while 26 samples were incorrectly classified. In addition, approximately 84.6% of the samples achieved an accuracy greater than 95% with mere 30,000 sequenced reads, corresponding to roughly 10 minutes of sequencing using two MinION flowcells or 20 minutes using one (**Fig. 4b**). Extending the sequencing to 150,000 reads resulted in 92.7% of samples reaching an accuracy above 95% (**Fig. 4b**).

Therefore, the integration of CNVisor and MethyLYZR significantly improves the accuracy of intraoperative glioma diagnostics, especially in the cases where read depth and sequencing quality are insufficient for a definitive diagnosis using either tool independently. In addition, this type of integration also reduces the minimum sequencing time needed to achieve accurate diagnosis. In summary, CNVs contain unique molecular information that is essential for accurately stratifying glioma subtypes. Incorporating CNVisor into the current methylation classifier such as MethyLYZR can significantly improve the accuracy and reduce the turnaround time of intraoperative diagnostics.

## Discussion

The utilization of CNV biomarkers to inform intraoperative decisions has been limited by the long turnaround time of conventional methods for genome-wide CNV characterization. Here, we have introduced CNVisor which enables accurate, genome-wide CNV analysis from ultra-low-coverage nanopore sequencing. CNVisor delivers probabilistic CNV profiles and provides a likelihood range for both chromosomal- and gene-level CNVs within tight surgical timelines. In our study, CNVisor reliably detected clinically relevant alterations, including 1p19q co-deletion, chromosome 7 amplification, chromosome 10 deletion, CDKN2A/B homozygous deletion, and EGFR amplification, with high concordance with the pathological diagnosis and a turnaround time within one hour, aligning with the timeframe of neuro-oncological procedures [31].

CNVisor addresses the limitations of existing CNV characterization methods for obtaining clinically relevant CNVs using nanopore sequencing in an operating room. Algorithms designed for short-read sequencing with abundant coverage [37] can detect CNVs at the kbps level, but often yield unreliable results when coverage is low [26, 33]. CNVisor instead infers CNV ranges to report chromosome- and gene-level alterations. This enables robust CNV calling under stringent time constraints without adopting additional molecular procedures [32, 33, 35].

In this paper, genome-wide CNV profiles, including clinically relevant CNVs, can be identified within 1 hour of biopsy, including 40 minutes of DNA extraction and rapid library preparation, and 20 minutes of computational analysis. This turnaround time reflects the implementation used in this study rather than an intrinsic limit of CNVisor. Since CNVisor resolves CNV status once sufficient reads are sequenced, further increases in sequencing throughput and basecalling efficiency could further reduce the turnaround time. In addition, improvements in DNA extraction and rapid library-preparation protocols could further shorten the workflow. Moreover, CNVisor can be readily integrated with established molecular procedures, such as SMURF-Seq and iSCORED, offering the potential to shorten the turnaround time even further [32, 33, 35].

The generated CNV profiles integrate naturally with complementary molecular profiles, such as methylation, for glioma stratification. CNVs and IDH mutation status are essential for distinguishing among glioblastoma, astrocytoma, and oligodendroglioma [17, 18]. Our results demonstrate that intraoperative CNV analysis can provide a unique, orthogonal dimension of molecular information that complements intraoperative methylation classifiers and IDH mutation status inference [25–27, 29, 36, 43]. Although current methylation-based classifiers excel at assigning tumors to broad tumor classes, they provide limited insight into tumor grading or genomic alterations associated with prognosis [23, 45], such as CDKN2A/B homozygous deletion. By quantifying alteration likelihoods for clinically relevant regions across the whole genome, CNVisor could confirm, refine, or even correct tumor type assignments from methylation-based classifiers and flag poor-prognosis biomarkers in time to optimize the surgical strategy.

CNVisor, together with methylation-based classifiers, offers intraoperative molecular insights beyond a diagnosis based on preoperative radiology. These complementary molecular profiles may help determine surgical strategies that balance the extent of resection with the risk of neurological damage. For instance, these profiles may be particularly valuable for patients with diffusely infiltrating gliomas. Rapid confirmation of an IDH-mutant, 1p19q co-deleted oligodendroglioma lacking CDKN2A/B homozygous deletion could support a more extensive resection strategy, given the associated favorable prognosis. Conversely, confirmation of a grade IV IDH-mutant astrocytoma with CDKN2A/B homozygous deletion may support a more conservative, function-preserving resection to minimize neurological damage and maintain the quality of the remaining life.

More work remains to be done to enable the adoption of CNVisor in routine intraoperative use. First, this paper is based on a limited number of cases. Our current implementation has not yet been validated or optimized for the intraoperative variability encountered in practice, such as differences between centers, instruments, and workflows. Variability in specimens, such as tumor purity and sequencing quality, needs to be systematically evaluated. Second, although we demonstrated robust detection of selected, clinically relevant CNVs across three major CNS tumor subtypes defined in current guidelines [17, 18], the clinical utility of the broader CNV landscape across all CNS tumor types remains incompletely characterized. CNVs can be associated with outcome or therapy response in specific contexts. Real-time interpretation of intraoperative CNV profiles of roughly 100 CNS tumor types [43] will require close collaboration with neuropathologists and molecular tumor boards. Third, and most importantly, no surgical management was altered based on the CNVisor output. This study demonstrates technical feasibility, but it does not yet establish decision impact or provide clinical benefit. Prospective clinical studies are required to evaluate CNVisor’s clinical utility and efficacy.

Despite these limitations, our work illustrates that accurate, genome-wide CNV profiles can be achieved within the timeframe of neurosurgery using nanopore sequencing, even under ultra-low coverage condition. Prospective, multicenter studies are essential to evaluate whether CNVisor can optimize surgical decisions and improve patient outcomes. The parameters and settings of CNVisor should be optimized and standardized for routine use. More broadly, CNVisor illustrates how real-time, probabilistic CNV analysis, together with methylation-based classifiers, can be integrated into neuro-oncology care. This paper supports a shift toward genomics-informed CNS tumor stratification, complementing conventional radiologic and histopathologic assessments.

## Materials and Methods

### Brain tumor cell line in vitro culture

Frozen cryovials of the brain tumor cell lines LN229 and U251MG were obtained from the Department of Neurosurgery, University Hospital Center Schleswig-Holstein. Thawed cells were cultured in T75 cell culture treated plastic flask in DMEM-F12 (Thermo Fisher Scientific) supplemented with 10% FCS and 1% penicillin/streptomycin. Once the cells have reached 90% confluency, they were passaged in a 1:5 ratio using 0,25% Trypsin/0.02% EDTA for expansion. Finally, cells were tested for the absence of Mycoplasma and subjected to DNA extraction and liquid nitrogen storage.

### Sample preparation

The DNA extraction from brain tumor tissue samples was done by taking advantage of the QIAamp Fast DNA tissue kit (Qiagen) with minor modification. Briefly 15 mg of tissue sample was transferred into a tissue disruption tube and supplemented with 265 µl digestion buffer. Tissue disruption was achieved by using a TissueLyser LT (Qiagen) with oscillation of 45 Hz for 2 min at room temperature. Disrupted tissue was immediately subjected to a proteinase K and RNase A digestion on a Thermomixer (Eppendorf) at 56°C for 7 min. The lysate was resuspended with 265 µl buffer MVL and loaded onto a QIAamp Mini spin column for DNA binding and subsequent washing steps. Purified DNA was finally eluted with 50 µl of preheated (56°C) nuclease-free H_2_O for 1 min and 2 µl of the eluate was quantified by using a Nanodrop One (Thermo Fisher Scientific). Extraction of DNA from brain tumor cell lines LN229 and U251MG was done using the GeneJET genomic DNA purification kit according to the manufacturers protocol. A modified phenol-chloroform extraction was used for the DNA extraction of brain tissue sample IEG4 according to a recently published protocol [46].

### Preparation of Nanopore sequencing libraries

Nanopore sequencing of DNA extracted from brain tumor tissue was performed using Oxford Nanopore Technology ligation sequencing kit (SQK-LSK109) and the rapid sequencing kit (SQK-RAD004) with minor modification to the manufacturer’s protocols. Briefly 2 x 6 µg of input DNA was used for preparation of two ligation sequencing libraries. End repair and dA-tailing (NEBNext Ultra II End Repair/dA-Tailing Module) was carried out at 20°C for 15 min following a 5 min heat-inactivation step at 65°C. The DNA was bound to Ampure XP beads (Beckman Coulter) for 5 min and washed twice with 80% Ethanol. Washed library was eluted for 20 min at 48°C and subjected to the ligation of ONT sequencing adapters (AMX). Ligation reaction was incubated for 30 min at room temperature following additional wash steps with long fragment buffer (LFB). Elution with ONT’s elution buffer (EB) was done for 20 min at 48 °C and up to 1,4 µg of final library was loaded onto a primed Promethion flow cell (FLO-PRO002). The flow cell was incubated for 20 min and sequencing was performed following two nuclease flush washes every 20-24 h. For preparation of rapid sequencing libraries up to 600 ng of DNA was used and subjected to the tagmentation reaction (FRA) according to the manufacturers protocol. The tagmented DNA was supplemented with 1 µl of rapid adapter (RAP) and incubated for 10 min at room temperature. The final library was loaded onto a primed Minion flow cell (R9.4.1) and sequenced for up to 72 h.

### Basecalling

The glioma samples and cell line samples were all basecalled using Guppy v6.3.8 (developed and provided by Oxford Nanopore Technologies). The surgical specimens were basecalled using Megalodon 2.5.0 and Guppy 6.0.6 (developed and provided by Oxford Nanopore Technologies). The configuration files used for basecalling can be found in Supplementary Table.

### Genome binning and mappability

We specifically used CHM13v2 reference genome [47] for improved alignment [48]. We partitioned each chromosome of the CHM13v2 reference genome into 5 Mbps bins and then evaluated whether the sampled DNA sequences of the reference genome can be mapped back to their original locations (Supplementary Document and Fig. S2a). After confirming that centromeres and telomeres are difficult to map regardless of the read length, we excluded centromeres and telomeres in CHM13v2 [47] for CNV characterization in this paper (Supplementary Document and Fig. S2b). Then we partitioned each chromosome arm of the CHM13v2 reference genome with centromeres and telomeres excluded into 3 Mbps bins for CNV characterization to balance between resolution and accuracy.

### Statistical model of CNVisor

Real-time sequencing may be conceptualized as a stochastic sampling of reads from the genome. The number of reads that can be sequenced in a clinical setting (less than 1 hour) are significantly smaller than the actual number of reads sequenced from a complete sequence run (up to 72 hours). Therefore, we can assume that the probability of obtaining reads from a given genomic region is not affected by the number of reads already obtained from this region. Additionally, we assume that a given genome of size *G* (human genome is 3 Gbps) is evenly split into *b* bins with each bin size of *S* = *G*⁄*b*. *L* is defined as the average read length. We assume that *S* ≫ *L* and that a read is assigned to a bin where the first base of the read is aligned to. Therefore, we can assume that each read aligns to one and only one bin.

Then the expected number of reads aligned to a normal diploid bin should be 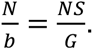 If we presume that a bin has a ploidy factor of *c* with actual CN of 2*c* (*c* = 1 for diploid, *c* > 1 for amplification, and *c* < 1 for deletion), then the expected number of reads aligned to this bin is given by 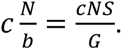 Then the probability of *n* out of *N* reads are aligned to this bin can be modeled as binomial distribution.

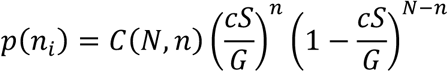

Assuming that we observe *n_i_* out of *N* reads aligned to a bin *B_i_*. The likelihood of observed *n_i_* under condition 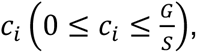 the ploidy factor of *B_i_*, is written as,

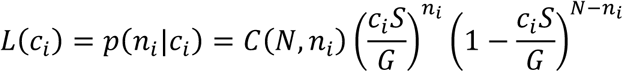

*L*(*c_i_*) can be normalized and rewritten in the form of Beta distribution (**Supplementary Document**),

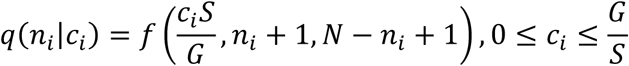

where 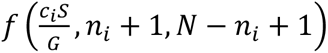 is the density function of Beta distribution [49]. We define 1 − *α* as the cumulative normalized likelihood *q*(*n_i_*|*c_i_*) between lower boundary *c*_min_ and upper boundary *c*_max_. i.e.,

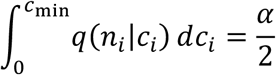

And

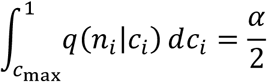

The boundaries can be obtained with quantiles from the beta distribution [50]:

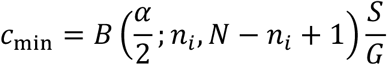

and

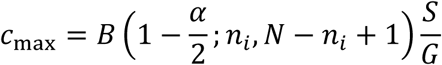

Since CNV can influence regions up to a whole chromosome, adjacent bins with similar CN values can be merged into the same segment. We assume that alignment processes to different bins are independent from each other. We want to obtain the probability that their ploidy factor difference is smaller than *Δ*.

Assume that *n_i_* and *n_i_*_+1_ reads are respectively observed to be aligned to adjacent bins *B_i_* and *B_i_*_+1_ of the same size. The bivariate likelihood that we observe such data under conditions 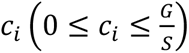 and 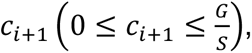 the ploidy factors of *B_i_* and *B_i_*_+1_ is given as the product of two independent binomials, i.e.,

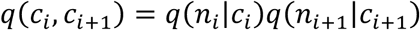

Then the cumulative bivariate likelihood that bins *B_i_* and *B_i_*_+1_ have ploidy factors smaller than *Δ* can be given as (Fig. S3d-f),

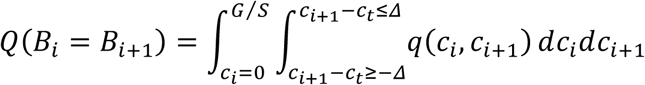

Similarly, the cumulative bivariate likelihoods that the CNV of *B_i_* smaller and larger than *B_i_*_+1_, *Q*(*B_i_* < *B_i_*_+1_) and *Q*(*B_i_* > *B_i_*_+1_) can also be obtained (**Supplementary Document**).The segmentation decision can be made based on the likelihood values of these three conditions, i.e., *Q*(*B_i_* = *B_i_*_+1_), *Q*(*B_i_* < *B_i_*_+1_), and *Q*(*B_i_* > *B_i_*_+1_). After the initial segmentation, the breakpoints are biased by the initial bin setting. We then generate the two bins next to each breakpoint into 5 sub-bins and optimize the breakpoints based on the read counts of these sub-bins using the same method described above. We used *Δ* of 0.15 in this paper.

### CNV characterization

The basecalled sequences were aligned to CHM13v2 reference genome using minimap2 v2.24 [51]. The unmapped reads, secondary alignments and supplementary alignments were removed using samtools [52] v1.17 with parameter -F 2308. The alignments with mapping quality lower than 10 were also removed (samtools parameter -q 10). Number of reads in each bin are counted using the alignment information and was normalized by the expected number of read to obtain the CNV values. Neighboring bins were merged into the same segment if they have CN differences less than Δ (Supplementary Document). In this paper, we set Δ = 0.15 everywhere. For each initial breakpoint, we further subdivided the two adjacent bins, one on each side of the breakpoint, into smaller sub-bins. The breakpoint position is optimized based on the read counts within these sub-bins. (Supplementary Document). The CNV range of each bin is estimated by likelihood function. The CNV range of each segment is adjusted by Bonferroni correction (Supplementary Document).

The likelihood of presence or absence of amplification or deletion are estimated (Supplementary Document). The likelihood of deletion or amplification in a bin or segment is quantified by the probability that the CNV value falls below -0.3 or exceeds 0.3, respectively. The likelihood of a bin or segment not being deleted or amplified is respectively given by the probabilities that the CNV value is larger or smaller than 0. 1p19q is co-deleted if the likelihood that more than 70% of each arm is deleted is larger than 50% (**Supplementary Document**). 1p19q is not co-deleted if the likelihood that more than 30% of either arm is not deleted is larger than 50% **(Supplementary Document)**. CDKN2A/B is considered to be deleted if its deletion likelihood is above 50% and not deleted if its likelihood of no deletion is above 50%. The status of chr7 amplification, chr10 deletion, and EGFR amplification are also decided using the same criteria. The CNV status is confirmed only when its alteration remains stable for 10 minutes.

### CNV characterization from Infinium MethylationEPIC BeadChips

The EZ DNA Methylation kit (ZymoResearch, Irvine, CA, USA) was used according to the manufacturer’s instructions to perform bisulfite conversion of the DNA. The converted DNA is then processed and hybridized to Infinium MethylationEPIC BeadChips (Illumina Inc., San Diego, CA, USA) following Illumina’s standard procedure. The chips are scanned using Illumina NextSeq550 system on default settings. CNV characterization of workflow is done using the circular binary segmentation algorithm included in the R package conumee. The results are visualized (CNV.genomeplot function with parameter detail=FALSE).

### Comparison between CNVisor and EPIC array

CNV profiles generated by CNVisor and those derived from EPIC array data were compared within a common genomic binning framework. The binning method used for CNVisor is described above. To make the two platforms directly comparable, each chromosome arm excluding the telomeres and centromere in the EPIC array CNV profile was divided into the same number of bins as in CNVisor. This created a shared set of genomic intervals.

For the EPIC array data, the CNV value for each bin was calculated as the weighted average of all EPIC array CNV segments overlapping with that bin, where the weights were given by the sizes of the overlaps. For CNVisor, the CNV value for each bin was taken directly from the CNVisor output. Bins on the sex chromosomes were excluded from the comparison.

Genome-wide similarity between the two binned CNV profiles was quantified using the Aitchison distance [41]. Since this metric is defined for positive values, CNV values were transformed to ploidy factors. To assess whether the observed similarity between CNVisor and EPIC array profiles was due to chance, we performed a permutation test in which the CNVisor bin values were randomly shuffled 10,000 times and the Aitchison distance was recalculated for each permutation. The observed distance was then compared with the null distribution for statistical significance.

### MethyLYZR

We downloaded and installed MethyLYZR from github (https://github.com/marasteiger/MethyLYZR). We used the recommended procedure [27]. The same reads used for CNV characterization of surgical specimens were applied to MethyLYZR.

### Sensitivity and specificity

Surgical specimen IEG63 and IEG127 with complete sequence runs were used to evaluate the sensitivity and specificity. Reads are subsampled from the complete datasets for each sample. 1p19q co-deletion and CDKN2A/B homozygous deletion are evaluated for each subsample reads. The sensitivity, or true positive rate, is represented by the ratio of correctly predicted alterations out of all the positive predicted events, i.e., the ratio of 1p19q co-deletion and CDKN2A/B homozygous deletion in subsampled IEG63. The specificity, or negative rate, given by the ratio of truly negative predicted alterations out of all negative events, i.e., the ratio of absence of 1p19q co-deletion nor CDKN2A/B homozygous deletion in subsampled IEG127.

### Subsampling of gliomas sequenced by R10 flowcells

We subsampled respectively 1,000 to 30,000 reads with an interval of 1,000, and 30,000 to 150,000 reads with an interval of 10,000. Each number of read number and each sample is subsampled 100 times.

### Integration of CNV and methylation profiles in glioma stratification

If the CNV profiles from CNVisor align with the classification from MethyLYZR, the glioma subtype is determined by the consensus of both methods. In cases where CNVisor and MethyLYZR disagree, the following process is applied. IDH mutation status is first inferred from MethyLYZR. For IDH mutated gliomas, the 1p19q co-deletion status from CNVisor is used to indicate the subtype, where the 1p19q co-deletion indicates oligodendroglioma, the absence 1p19q co-deletion indicates astrocytoma, and the inconclusive 1p19q status indicates inconclusive glioma subtype. For IDH wild-type glioma with low MethyLYZR confidence (lower than 0.5), the glioma subtype is inferred from CNVisor profiles. For IDH wild-type gliomas with high confidence in MethyLYZR (higher than 0.5), the status of chromosomes 7 and 10, as well as EGFR, is used to determine glioblastoma. If these do not indicate glioblastoma, the subtype is determined solely by MethyLYZR.

## Data availability

The human telomere-to-telomere reference genome is available from github (https://github.com/marbl/CHM13/blob/master/Sequencing_data.md). The trio data are available from Amazon Web Server (s3://ont-open-data/giab_lsk114_2022.12). Requests of academic use of nanopore sequencing data from cell lines and glioma samples can be upon request to the corresponding author.

## Code availability

CNVisor is available on Github: https://github.com/GJYWang/CNVisor.git. The visualization used in this paper is available on Github: https://github.com/GJYWang/CNVisor_wg_visual.

## Data Availability

All data produced in the present study are available upon reasonable request to the authors

## Acknowledgement

We are deeply thankful for the invaluable support of the patients and their families who graciously donated biomaterials for this study. We also thank the IT department of the Max Planck Institute for Molecular Genetics for assistance with data management.

M.E. was supported by the Bundesministerium für Bildung und Forschung (BMBF, German Federal Ministry of Education and Research) (IntraEpiGliom, FKZ 13GW0347A). C.R. was supported by the BMBF (IntraEpiGliom, FKZ 13GW0347B). B.B., M. Steiger, R.S., G.W. and F.-J.M. were supported by the BMBF (IntraEpiGliom, FKZ 13GW0347C). C.K. and M. Synowitz were supported by the BMBF (IntraEpiGliom, FKZ 13GW0347D). F.-J.M. was supported by the BMBF (P4D, FKZ 01EK2204C). B.B. and F.-J.M. were funded by the Deutsche Forschungsgemeinschaft (DFG, German Research Foundation) under Germany’s Excellence Strategy – EXC 2167-390884018 (Cluster of Excellence “Precision Medicine in Chronic Inflammation”, PMI) and EXC 2167/2 (PMI, second funding period) and CRC 1665 – 515637292, and by an intramural grant from the University Comprehensive Cancer Center Schleswig-Holstein (UCCSH Twinning 2022). M. Steiger and H.K. were supported by the Max Planck Society. S.Y. is supported by the Health Professional Investigator Award (HPI-2022-2834) from Michael Smith Health Research BC.

## Author contribution

G.W., M.V. and F.J.M. conceived the study and designed the experiments. G.W. and M.V. developed the mathematical model. G.W. conducted the statistical analysis. G.W. and K.Z. implemented the computational pipeline. G.W., K.Z., and R.S. performed the analyses. B.B. sequenced the cell line and glioma samples. G.W. and C.R. conducted data processing. D.W., A.L., and S.Y. provided patient glioma samples and molecular information. G.W., R.S., M.V. and F.J.M. analyzed the results. All authors wrote the manuscript. M.V. and F.J.M. supervised the research.

## Competing interests

S.Y. is a member of the advisory board and has received honoraria from Amgen, AstraZeneca, Bayer, Pfizer, Roche, and Servier. M.V. is a shareholder and member of the scientific advisory board of Lucid Genomics.

---

**Fig. S1.**
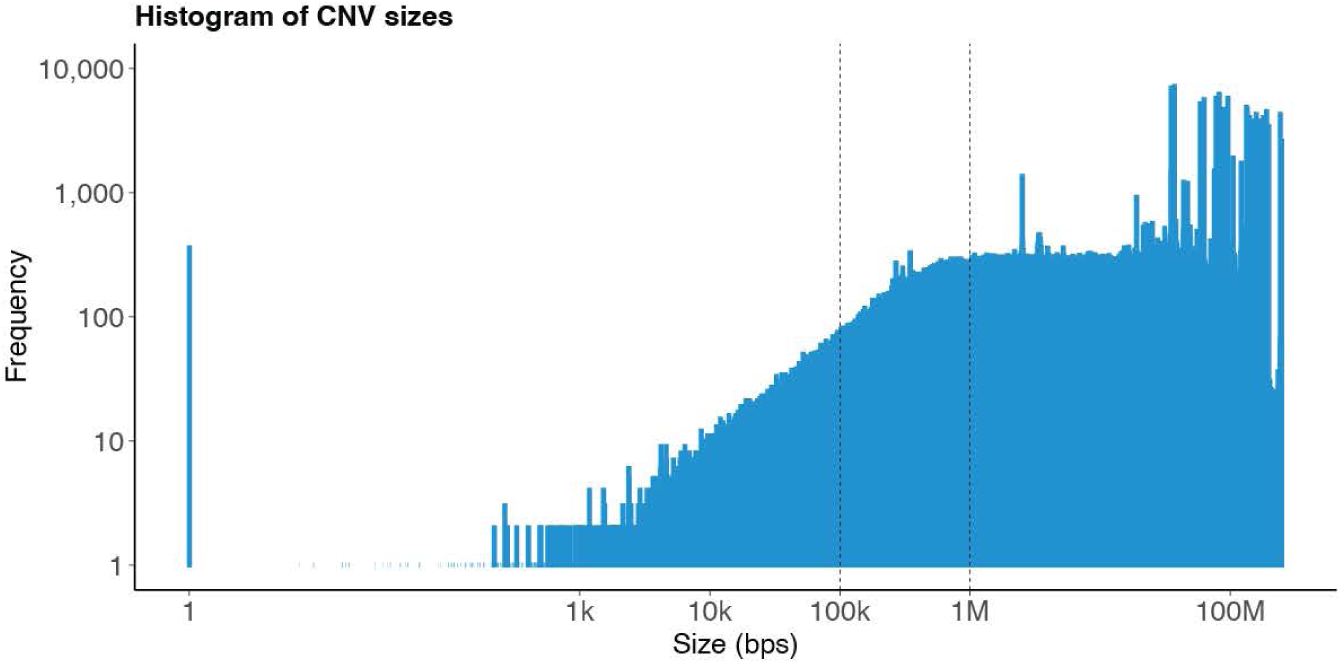
The histogram of the sizes of CNV signatures obtained from TCGA. The x-axis is the sizes of the CNVs and the y-axis is the counts. More than 97% of the CNVs have sizes larger than 100 kbps and more than 78% of the CNVs have sizes larger than 1 Mbps.

**Fig. S2.**
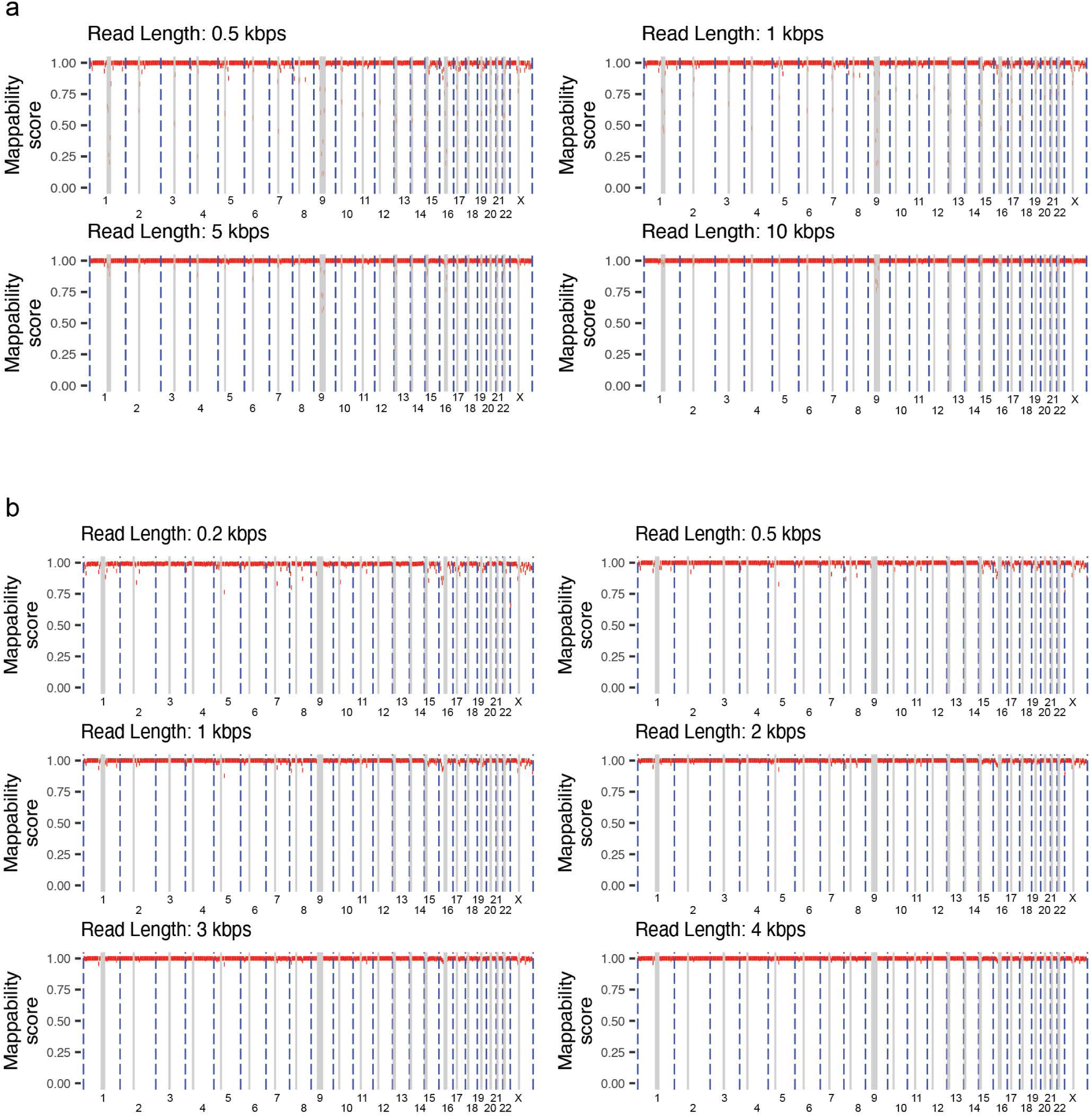
Mappability score of CHM13v2 with different read lengths. The score is given by the percentage of reads sampled from this region that can be mapped back to their original locations (Supplementary Document). X-axis represents the genome and y-axis represents mappability score. (a) The mappability score of whole CHM13v2 using a read length of 0.5/1/5/10 kbps sampled with an interval of 20/50/250/250 bps. (b) The mappability score of CHM13v2 excluding telomere and centromeres using a read length of 0.2/0.5/1/2/3/4 kbps with an interval of 5/20/50/100/150/200 bps.

**Fig. S3.**
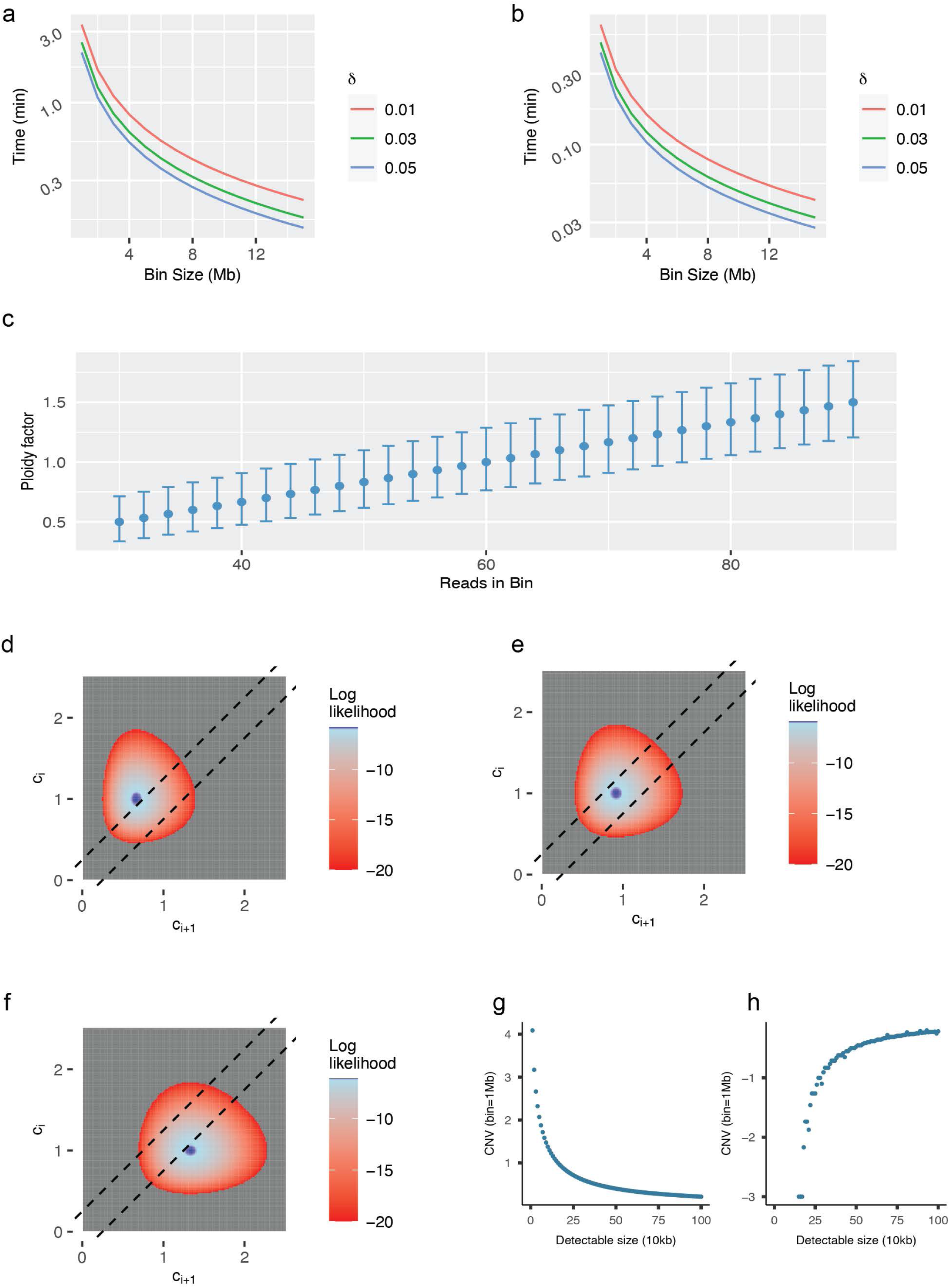
Statistical analysis of real-time CNV characterization. (a) Minimum time required so that less than *δ*(0 ≤ *δ* ≤ 1) of the genome is not covered by any read with different bin sizes using a MinION in an ideal scenario. (b) Minimum time required so that less than *δ*(0 ≤ *δ* ≤ 1) of the genome is not covered by any read with different bin sizes using a PromethION in an ideal scenario. (c) Simulation of the CN and its range with different number of aligned reads when 60,000 reads are sequenced and the genome is evenly split into bins of 3 Mbps. (d)-(f) The log likelihood (gray area is log likelihood less than -20) of that two bins have CN difference smaller than *Δ* = 0.3 if 60 reads are aligned to *B_i_* and 40, 55, and 80 reads are aligned to *B_i_*_+1_ with 60,000 reads sequenced and the genome is split into 1000 bins. The cumulative likelihood of region above the top line represents the likelihood that *B_i_*_+1_ is smaller than *B_i_*. The cumulative likelihood of region between two lines represents the likelihood that *B_i_*_+1_ and *B_i_* have differences less than *Δ*. The cumulative likelihood of region below the bottom line represents the likelihood that *B_i_*_+1_ is larger than *B_i_*. (g)-(h) Simulation of the detectable CNVs, assuming that 150,000 reads are sequenced (average sequenced reads of two MinION flowcells in 20 minutes) and the genome is evenly split into bins of 1 Mbps. Assuming that a bin adjacent to a diploidy bin contains CNVs smaller than the bin size. The relationship between CNV sizes and the minimum amplification that can be detected (d). The relationship between CNV sizes and the maximum deletion that can be detected (d).

**Fig. S4.**
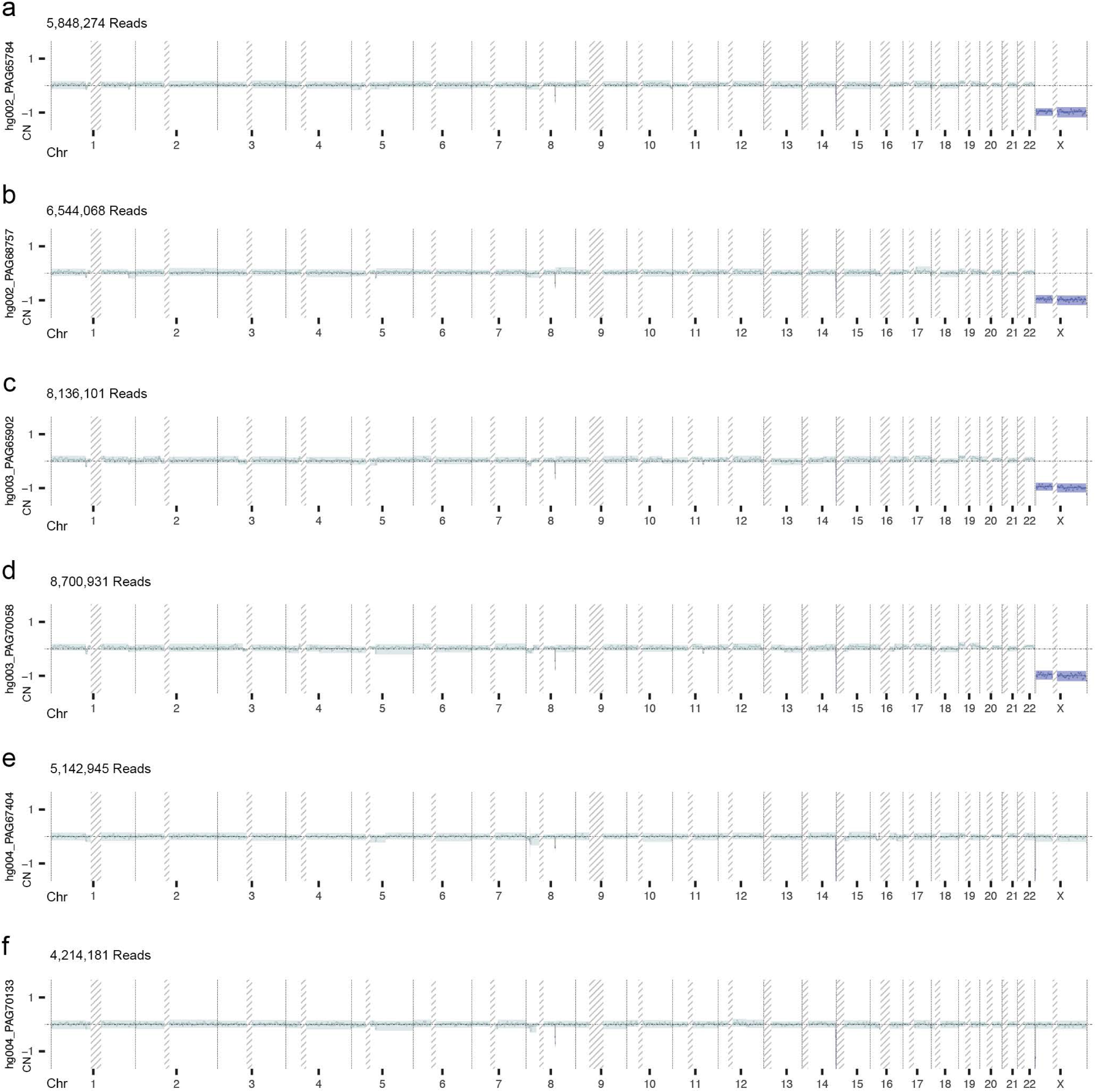
CNV characterization of Ashkenazi Trio data using CNVisor. (a) HG002 sequenced with flowcell PAG65784. (b) HG002 sequenced with flowcell PAG68757. (c) HG003 sequenced with flowcell PAG65902. (d) HG003 sequenced with flowcell PAG70058. (e) HG004 sequenced with flowcell PAG67404. (f) HG004 sequenced with flowcell PAG70133. X-axis represents the genome and y-axis represents log2 ratio CN. Blue, red, and light green respectively represent amplification, deletion, and no alteration. The colored line is the CNV of the bin/segment, and the colored shade is its CNV range.

**Fig. S5.**
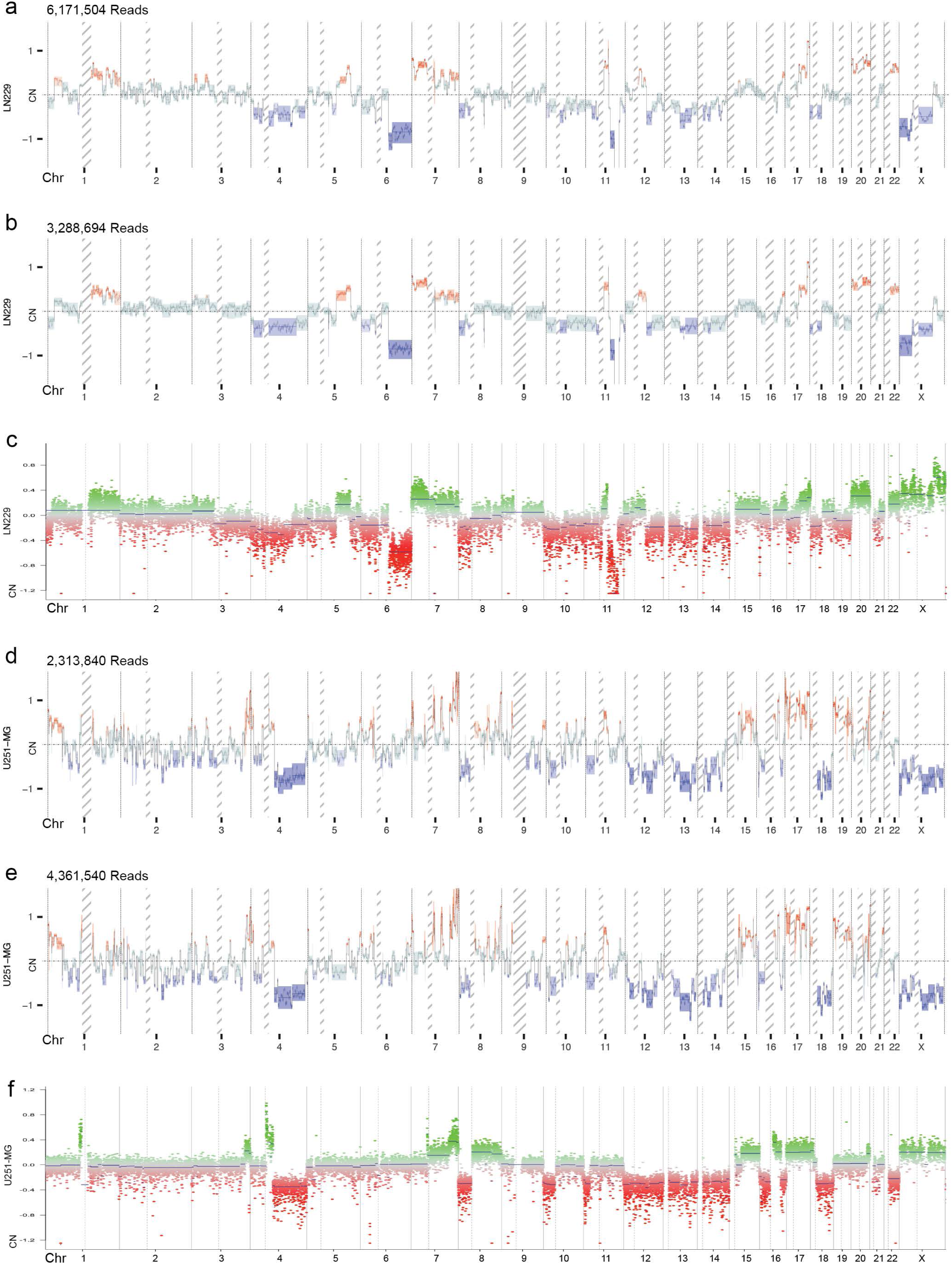
CNV characterization of glioma cell line using CNVisor and EPIC array. (a) Glioma cell line LN229 sequenced with flowcell PAD70703. (b) Glioma cell line LN229 sequenced with flowcell FAM96122. (c) Glioma cell line LN229 characterized by EPIC array. (d) Glioma cell line U251-MG sequenced with flowcell PAD15032. (e) Glioma cell line U251-MG sequenced with flowcell PAD70709. (f) Glioma cell line U251-MG characterized by EPIC array. In (a), (b), (d), and (e), X-axis represents the genome and y-axis represents log2 ratio CN. Blue, red, and light green respectively represent amplification, deletion, and no alteration. The colored line is the CNV of the bin/segment, and the colored shade is its CNV range. In In (c), and (e), X-axis represents the genome and y-axis represents log2 ratio CN. Green, and red respectively represent CN larger and smaller than 0.

**Fig. S6.**
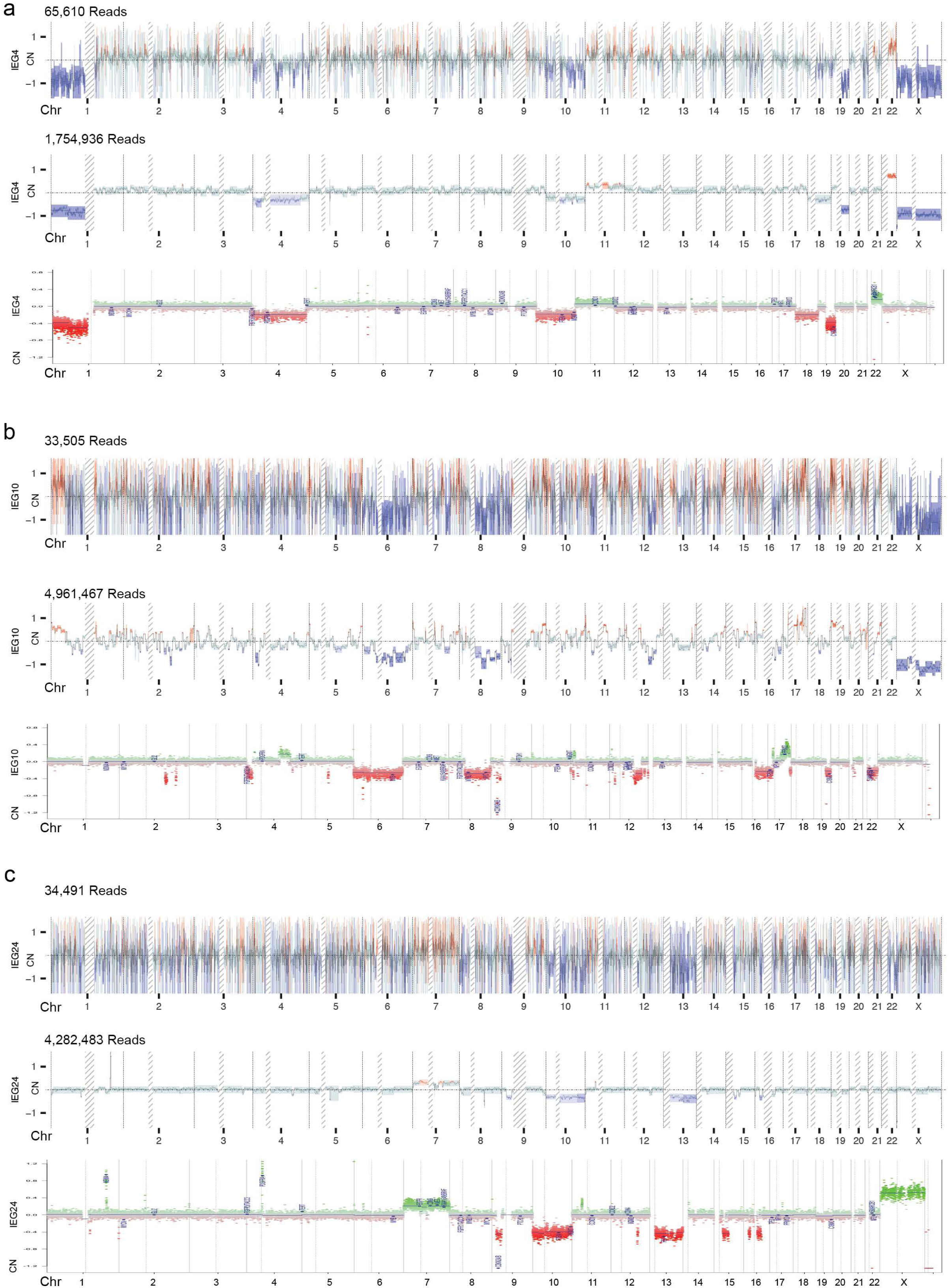
Real-time simulation of CNV characterization at 60 minutes of sequencing using glioma samples with known diagnosis. (a) IEG4 sequenced with flowcell PAD26308. (b) IEG10 sequenced with flowcell PAD70822. (c) IEG24 sequenced with flowcell FAO43670. X-axis represents the genome and y-axis represents log2 ratio CN. Blue, red, and light green respectively represent amplification, deletion, and no alteration. The colored line is the CNV of the bin/segment, and the colored shade is its CNV range.

**Fig. S7.**
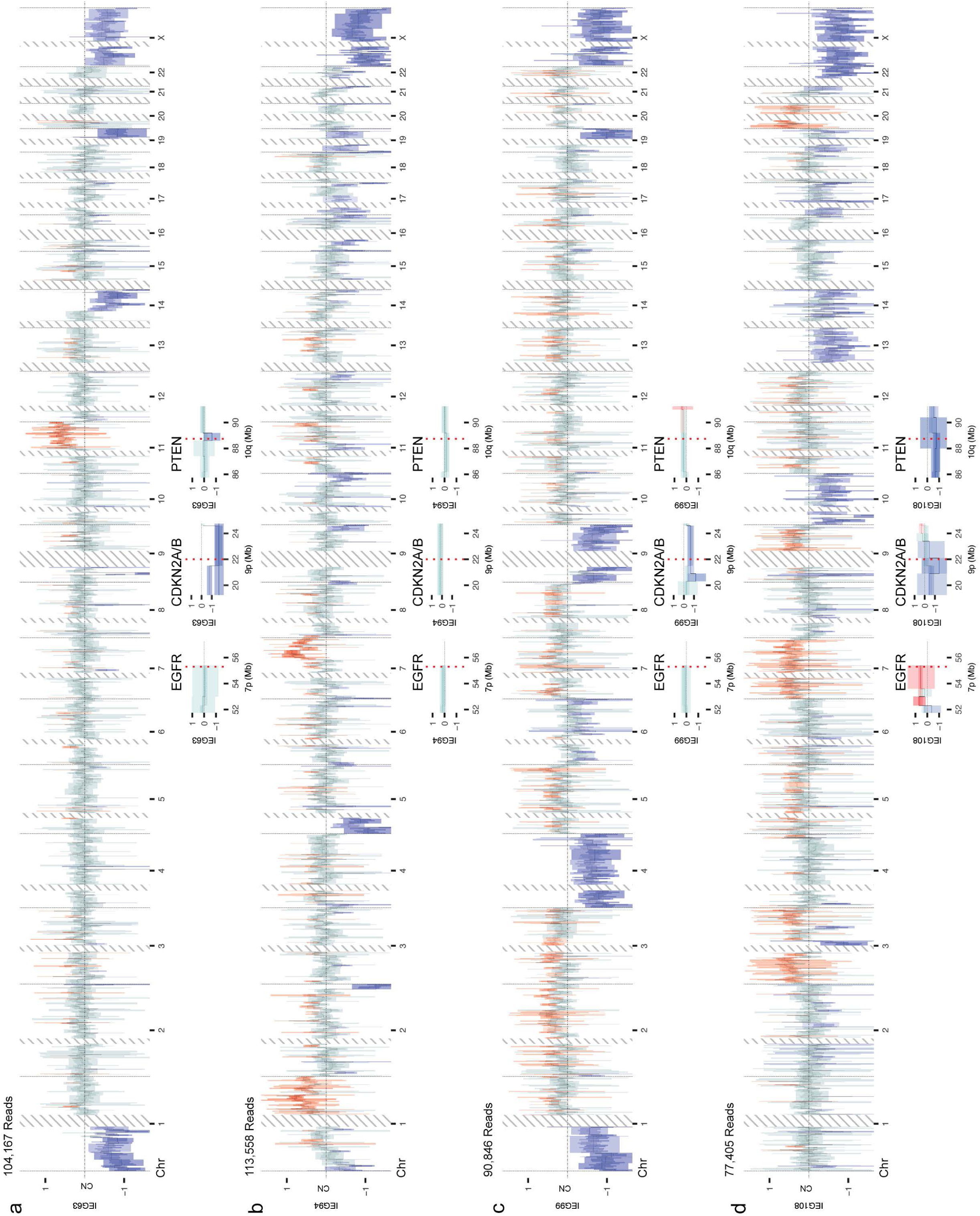
Real-time CNV characterization of surgical specimens at 30 minutes of sequencing using multiple flowcells. (a) IEG63 sequenced with flowcells FAP22594 and FAO65859. (b) IEG94 sequenced with flowcells FAP54529 and FAP55076. (c) IEG99 sequenced with flowcells FAO98209 and FAO61374. (d) IEG108 sequenced with flowcells FAQ67156, FAQ67431 and FAQ72189. X-axis represents the genome and y-axis represents log2 ratio CN. Blue, red, and light green respectively represent amplification, deletion, and no alteration. The colored line is the CNV of the bin/segment, and the colored shade is its CNV range.

**Fig. S8.**
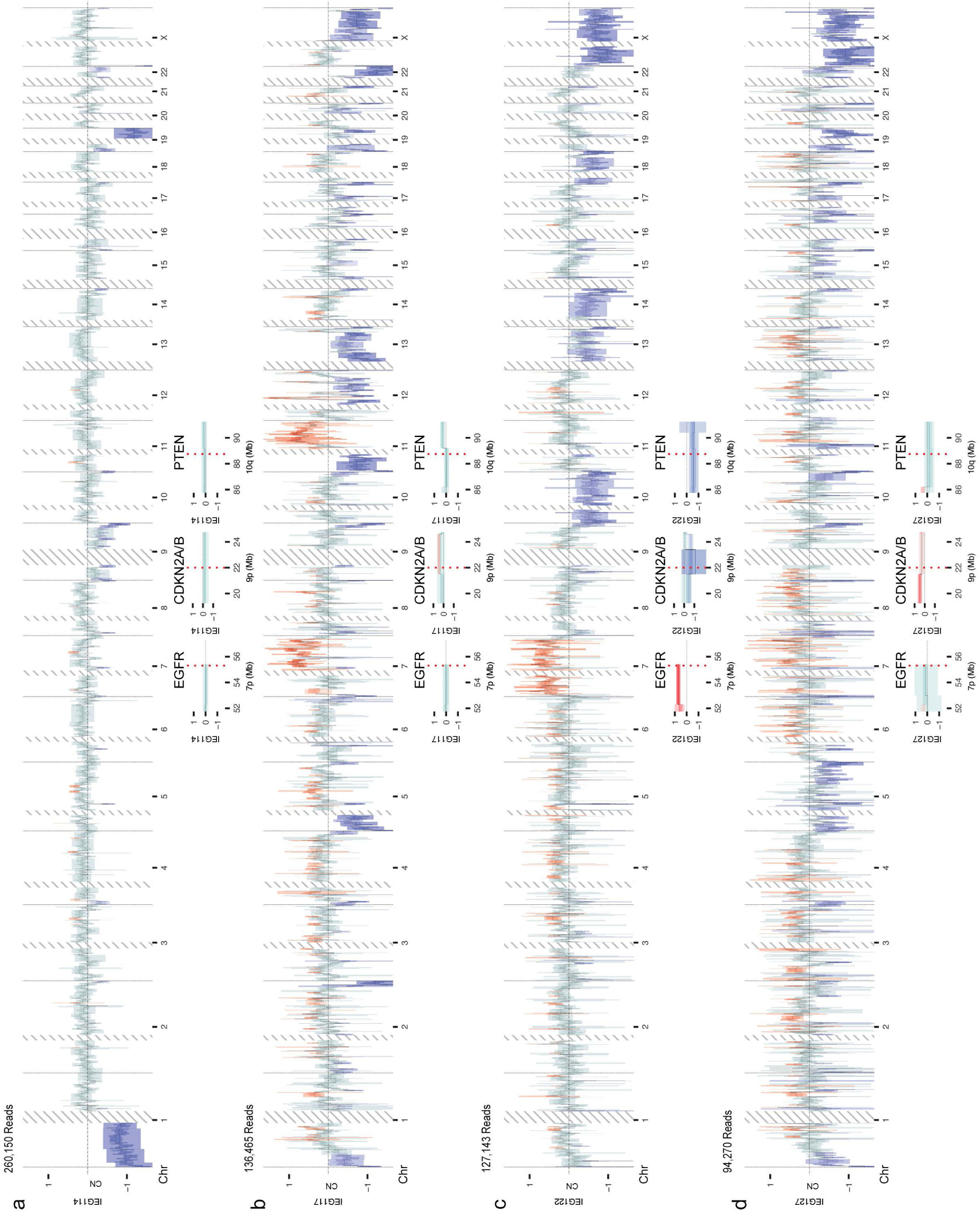
Real-time CNV characterization of surgical specimens at 30 minutes of sequencing using multiple flowcells. (a) IEG114 sequenced with flowcells FAQ67345, FAQ68170, FAQ67672 and FAQ67742. (b) IEG117 sequenced with flowcells FAR18179 and FAR18036. (c) IEG122 sequenced with flowcells FAR83238 and FAR83976. (d) IEG127 sequenced with flowcells FAR83224 and FAR83120. X-axis represents the genome and y-axis represents log2 ratio CN. Blue, red, and light green respectively represent amplification, deletion, and no alteration. The colored line is the CNV of the bin/segment, and the colored shade is its CNV range.

**Fig. S9.**
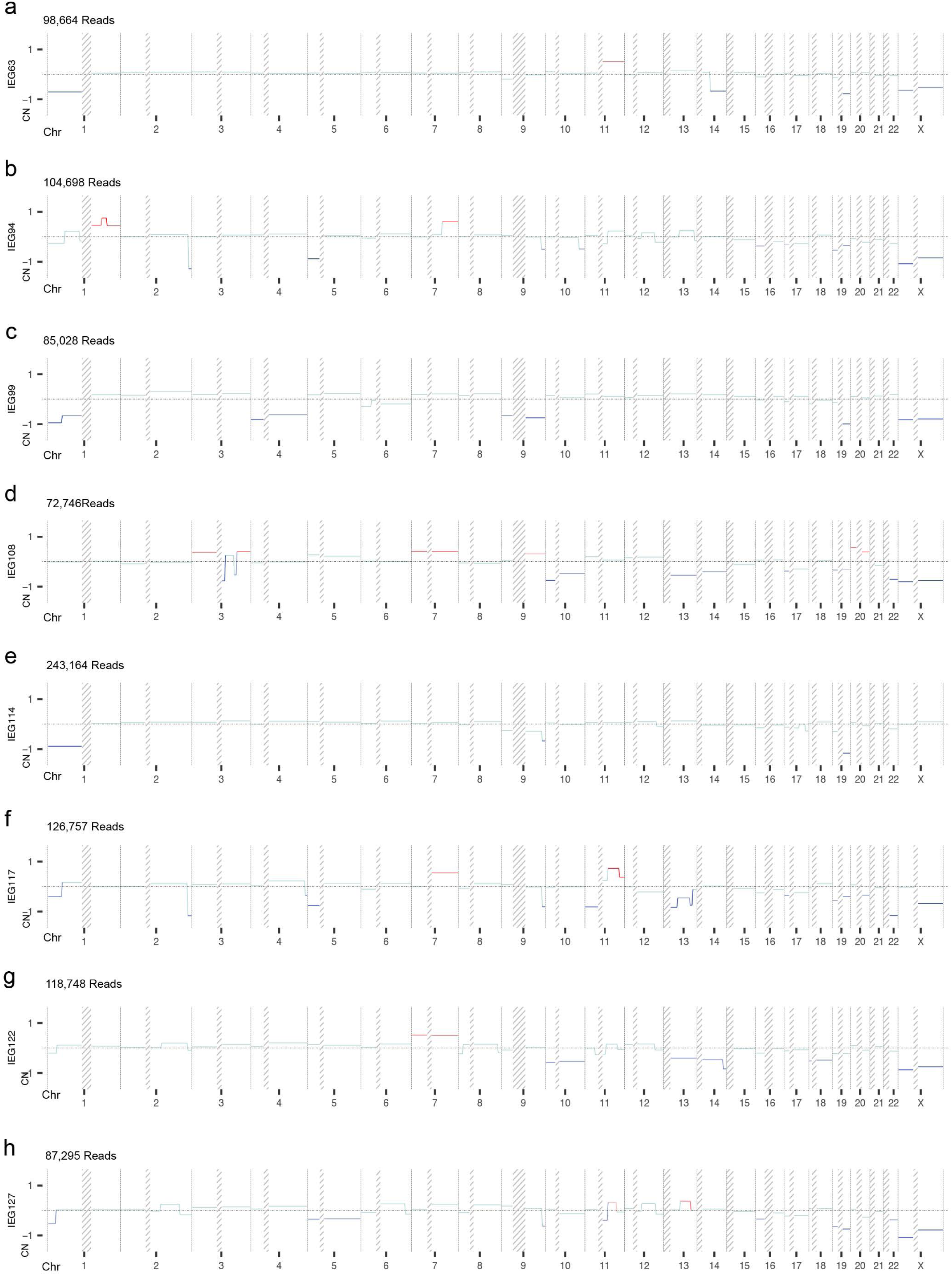
CNV profiles generated by Circular Binary Segmentation with surgical specimens using 30 minutes of sequencing from multiple flowcells (the exact same amount of data to generate Fig. 2, S7 and S8). (a) IEG63 sequenced with flowcells FAP22594 and FAO65859. (b) IEG94 sequenced with flowcells FAP54529 and FAP55076. (c) IEG99 sequenced with flowcells FAO98209 and FAO61374. (d) IEG108 sequenced with flowcells FAQ67156, FAQ67431 and FAQ72189. (a) IEG114 sequenced with flowcells FAQ67345, FAQ68170, FAQ67672 and FAQ67742. (b) IEG117 sequenced with flowcells FAR18179 and FAR18036. (c) IEG122 sequenced with flowcells FAR83238 and FAR83976. (d) IEG127 sequenced with flowcells FAR83224 and FAR83120. X-axis represents the genome and y-axis represents log2 ratio CN. Blue, red, and light green respectively represent amplification, deletion, and no alteration. The colored line is the CNV of the segment from Circular Binary Segmentation.

## Notes

### Author Declarations

The study protocol was approved by and adhered to the Clinical Ethics Committee of the Medical Faculty of Kiel University (D443/20). All included patients or their legal guardians/parents provided written informed consent for participation in the study. The results were not shared with treating physicians or caregivers and, therefore, not used to alter patient treatment or diagnosis. The study was approved by and adhered to the University of British Columbia Research Ethics Committee (REB no. H08-02838). The study was approved by and adhered to the Clinical Ethics Committee of the Medical Faculty of Regensburg University (20-1799-101).

## Reference

1. Hanahan, D. and Robert A. Weinberg, Hallmarks of Cancer: The Next Generation. Cell, 2011. 144(5): p. 646–674.

2. Hanahan, D., Hallmarks of Cancer: New Dimensions. Cancer Discovery, 2022. 12(1): p. 31–46.

3. Tang, Y.-C. and A. Amon, Gene Copy-Number Alterations: A Cost-Benefit Analysis. Cell, 2013. 152(3): p. 394–405.

4. Hastings, P.J., et al., Mechanisms of change in gene copy number. Nature Reviews Genetics, 2009. 10(8): p. 551–564.

5. Pinkel, D., et al., High resolution analysis of DNA copy number variation using comparative genomic hybridization to microarrays. Nature Genetics, 1998. 20(2): p. 207–211.

6. Winchester, L., C. Yau, and J. Ragoussis, Comparing CNV detection methods for SNP arrays. Briefings in Functional Genomics, 2009. 8(5): p. 353–366.

7. Chiang, D.Y., et al., High-resolution mapping of copy-number alterations with massively parallel sequencing. Nature Methods, 2009. 6(1): p. 99–103.

8. Baslan, T., et al., Genome-wide copy number analysis of single cells. Nature Protocols, 2012. 7(6): p. 1024–1041.

9. Navin, N., et al., Tumour evolution inferred by single-cell sequencing. Nature, 2011. 472(7341): p. 90–94.

10. Beroukhim, R., et al., Assessing the significance of chromosomal aberrations in cancer: Methodology and application to glioma. Proceedings of the National Academy of Sciences, 2007. 104(50): p. 20007–20012.

11. Zack, T.I., et al., Pan-cancer patterns of somatic copy number alteration. Nature Genetics, 2013. 45(10): p. 1134–1140.

12. Taylor, A.M., et al., Genomic and Functional Approaches to Understanding Cancer Aneuploidy. Cancer Cell, 2018. 33(4): p. 676–689.e3.

13. Berger, A.C., et al., A Comprehensive Pan-Cancer Molecular Study of Gynecologic and Breast Cancers. Cancer Cell, 2018. 33(4): p. 690–705.e9.

14. Campbell, J.D., et al., Genomic, Pathway Network, and Immunologic Features Distinguishing Squamous Carcinomas. Cell Reports, 2018. 23(1): p. 194–212.e6.

15. Steele, C.D., et al., Signatures of copy number alterations in human cancer. Nature, 2022. 606(7916): p. 984–991.

16. Luchini, C., et al., Molecular Tumor Boards in Clinical Practice. Trends in Cancer, 2020. 6(9): p. 738–744.

17. Louis, D.N., et al., The 2021 WHO Classification of Tumors of the Central Nervous System: a summary. Neuro Oncol, 2021. 23(8): p. 1231–1251.

18. Weller, M., et al., EANO guidelines on the diagnosis and treatment of diffuse gliomas of adulthood. Nature Reviews Clinical Oncology, 2021. 18(3): p. 170–186.

19. Comprehensive, Integrative Genomic Analysis of Diffuse Lower-Grade Gliomas. New England Journal of Medicine, 2015. 372(26): p. 2481–2498.

20. Brennan, Cameron W., et al., The Somatic Genomic Landscape of Glioblastoma. Cell, 2013. 155(2): p. 462–477.

21. McLendon, R., et al., Comprehensive genomic characterization defines human glioblastoma genes and core pathways. Nature, 2008. 455(7216): p. 1061–1068.

22. Brat, D.J., et al., cIMPACT-NOW update 3: recommended diagnostic criteria for “Diffuse astrocytic glioma, IDH-wildtype, with molecular features of glioblastoma, WHO grade IV”. Acta Neuropathologica, 2018. 136(5): p. 805–810.

23. Appay, R., et al., CDKN2A homozygous deletion is a strong adverse prognosis factor in diffuse malignant IDH-mutant gliomas. Neuro-Oncology, 2019. 21(12): p. 1519–1528.

24. Wang, Y., et al., Nanopore sequencing technology, bioinformatics and applications. Nature Biotechnology, 2021. 39(11): p. 1348–1365.

25. Kuschel, L.P., et al., Robust methylation-based classification of brain tumours using nanopore sequencing. Neuropathol Appl Neurobiol, 2023. 49(1): p. e12856.

26. Vermeulen, C., et al., Ultra-fast deep-learned CNS tumour classification during surgery. Nature, 2023. 622(7984): p. 842–849.

27. Brändl, B., et al., Rapid brain tumor classification from sparse epigenomic data. Nature Medicine, 2025. 31(3): p. 840–848.

28. Deacon, S., et al., ROBIN: A unified nanopore-based assay integrating intraoperative methylome classification and next-day comprehensive profiling for ultra-rapid tumor diagnosis. Neuro-Oncology, 2025. 27(8): p. 2035–2046.

29. Patel, A., et al., Prospective, multicenter validation of a platform for rapid molecular profiling of central nervous system tumors. Nature Medicine, 2025. 31(5): p. 1567–1577.

30. Magi, A., et al., Nano-GLADIATOR: real-time detection of copy number alterations from nanopore sequencing data. Bioinformatics, 2019. 35(21): p. 4213–4221.

31. Senders, J.T., et al., Thirty-Day Outcomes After Craniotomy for Primary Malignant Brain Tumors: A National Surgical Quality Improvement Program Analysis. Neurosurgery, 2018. 83(6): p. 1249–1259.

32. Baslan, T., et al., High resolution copy number inference in cancer using short-molecule nanopore sequencing. Nucleic Acids Research, 2021. 49(21): p. e124–e124.

33. Prabakar, R.K., et al., SMURF-seq: efficient copy number profiling on long-read sequencers. Genome Biology, 2019. 20(1): p. 134.

34. Wongsurawat, T., et al., Exploiting nanopore sequencing for characterization and grading of IDH-mutant gliomas. Brain Pathology, 2024. 34(1): p. e13203.

35. Emiliani, F.E., et al., Nanopore-based random genomic sampling for intraoperative molecular diagnosis. Genome Medicine, 2025. 17(1): p. 6.

36. Djirackor, L., et al., Intraoperative DNA methylation classification of brain tumors impacts neurosurgical strategy. Neuro-Oncology Advances, 2021. 3(1).

37. Olshen, A.B., et al., Circular binary segmentation for the analysis of array-based DNA copy number data. Biostatistics, 2004. 5(4): p. 557–572.

38. Gorzynski, J.E., et al., Ultrarapid Nanopore Genome Sequencing in a Critical Care Setting. N Engl J Med, 2022. 386(7): p. 700–702.

39. Patel, A., et al., Rapid-CNS2: rapid comprehensive adaptive nanopore-sequencing of CNS tumors, a proof-of-concept study. Acta Neuropathologica, 2022. 143(5): p. 609–612.

40. Shumate, A., et al., Assembly and annotation of an Ashkenazi human reference genome. Genome Biology, 2020. 21(1): p. 129.

41. Kerbs, P., et al., Employing nanopore sequencing on FFPE-derived DNA for CNS tumor diagnostics. Acta Neuropathologica Communications, 2025. 13(1): p. 226.

42. Schnöller, L.E., et al., Systematic in vitro analysis of therapy resistance in glioblastoma cell lines by integration of clonogenic survival data with multi-level molecular data. Radiation Oncology, 2023. 18(1): p. 51.

43. Capper, D., et al., DNA methylation-based classification of central nervous system tumours. Nature, 2018. 555(7697): p. 469–474.

44. Ceccarelli, M., et al., Molecular Profiling Reveals Biologically Discrete Subsets and Pathways of Progression in Diffuse Glioma. Cell, 2016. 164(3): p. 550–563.

45. Capper, D., et al., Practical implementation of DNA methylation and copy-number-based CNS tumor diagnostics: the Heidelberg experience. Acta Neuropathologica, 2018. 136(2): p. 181–210.

46. Sambrook, J., E.F. Fritsch, and T. Maniatis, Molecular cloning: a laboratory manual. 1989: Cold spring harbor laboratory press.

47. Nurk, S., et al., The complete sequence of a human genome. Science, 2022. 376(6588): p. 44–53.

48. Aganezov, S., et al., A complete reference genome improves analysis of human genetic variation. Science. 376(6588): p. eabl3533.

49. Gupta, A.K. and S. Nadarajah, Handbook of beta distribution and its applications. 2004: CRC press.

50. Måns, T., The cost of using exact confidence intervals for a binomial proportion. Electronic Journal of Statistics, 2014. 8(1): p. 817–840.

51. Li, H., Minimap2: pairwise alignment for nucleotide sequences. Bioinformatics, 2018. 34(18): p. 3094–3100.

52. Li, H., et al., The Sequence Alignment/Map format and SAMtools. Bioinformatics, 2009. 25(16): p. 2078–2079.

