## Supplementary Material for "Intraoperative copy number profiling from ultra-low coverage long-read sequencing for molecular tumor assessment"

* Equal contribution

**Section 1. Statistical analysis of real-time CNV characterization** **3**
**Section 2. Mappability and binning the genome** **9**
**Section 3. Coverage comparison between CNV characterization using NGS and nanopore sequencing** **10**
**Section 4. Comparison of segmentation methods** **11**

### Section 1. Statistical analysis of real-time CNV characterization

Real-time sequencing may be conceptualized as a stochastic sampling of reads from the genome. The number of reads that can be sequenced in a clinical setting (less than 1 hour) are significantly smaller than the actual number of reads sequenced from a complete sequence run (up to 72 hours). Therefore, we can assume that the probability of obtaining reads from a given genomic region is not affected by the number of reads already obtained from this region. Additionally, we assume that a given genome of size $G$ (human genome is 3 Gbps) is evenly split into $b$ bins with each bin size of $S = G/b$. $L$ is defined as the average read length. We assume that $S \gg L$ and that a read is assigned to a bin where the first base of the read is aligned to. Therefore, we can assume that each read aligns to one and only one bin.

In the analysis below, we use $N$ to represent the number of sequenced reads. Given the channel number $H$, sequencing speed $q$, and the pore occupancy rate $o$, the sequencing time of $N$ reads is $T = \frac{NL}{Hqo}$. Usually, a MinION has 512 channels and a PromethION has 2675. We assume that both sequencers can reach 90% of pore occupancy and 450bps/s sequencing speed per channel and that the average read length is 5kbps. These parameters will be used to estimate the required time in real-time sequencing. Additionally, we used log2 ratio to denote CNV values in this paper.

#### 1.1. Minimum time required for CNV characterization

We need to sequence long enough to have minimal 1 read in most of the bins. Assume that $B_i$ is a random bin in the genome. The probability that one or more reads aligned to a random base in $B_i$ is $\frac{N}{G}$. Since there are $S$ bases in $B_i$, the probability that no read is aligned to $B_i$ is given by

$$P(r_1 \dots r_N \notin B_i) = \left(1 - \frac{N}{G}\right)^S$$

Using Euler formula, this probability can be approximated to

$$P(r_1 \dots r_N \notin B_i) = \left(1 - \frac{N}{G}\right)^{\frac{G}{b}} = \left[\left(1 - \frac{N}{G}\right)^G\right]^{1/b} \approx e^{\frac{-N}{b}}$$

Define $\delta (0 \le \delta \le 1)$ as the upper limit of the fraction of bins without any aligned read, we can write this as,

$$P(r_1 \dots r_N \notin B_i) < \delta$$

Solving the equation gives:

$$N > -bln(\delta)$$

Then with a bin size of 5Mbps, a MinION needs to sequence for 1 minute so that less than 1% of the genome remains uncovered by any read while a PromethION only needs 15 seconds (Fig. S3a-b). Based on different assumptions, the minimum time required for CNV characterization can be estimated accordingly.

### 1.2. CNV range of a bin

The expected number of reads aligned to a normal diploid bin should be $\frac{N}{b} = \frac{NS}{G}$. If we presume that a bin has a ploidy factor of $c$ with actual CN of $2c$ ($c = 1$ for diploid, $c > 1$ for amplification, and $c < 1$ for deletion), then the expected number of reads aligned to this bin is given by $c\frac{N}{b} = \frac{cNS}{G}$. Then the probability of $n$ out of $N$ reads are aligned to this bin can be modeled as binomial distribution.

$$p(n_i) = C(N, n)\left(\frac{cS}{G}\right)^n \left(1 - \frac{cS}{G}\right)^{N-n}$$

Assuming that we observe $n_i$ out of $N$ reads aligned to a bin $B_i$. We want to obtain the likelihood of observed $n_i$ under condition $c_i\left(0 \le c_i \le \frac{G}{S}\right)$, the ploidy factor of $B_i$. $c_i$ is continuous since we are sequencing a mixture of cancer and normal cells and $N \gg 1$. The likelihood function is written as,

$$L(c_i) = p(n_i|c_i) = C(N, n_i)\left(\frac{c_iS}{G}\right)^{n_i} \left(1 - \frac{c_iS}{G}\right)^{N-n_i}$$

where

$$C(N, n_i) = \frac{N!}{n_i!\,(N - n_i)!} = \frac{\Gamma(N+1)}{\Gamma(n_i+1)\Gamma(N-n_i+1)} = \frac{\Gamma\,(N+2)/(N+1)}{\Gamma(n_i+1)\Gamma(N-n_i+1)}$$

$L(c_i)$ can be rewritten as

$$L(c_i) = \frac{1}{N+1} f\left(\frac{c_iS}{G}, n_i + 1, N - n_i + 1\right)$$

where $f(t, n_i + 1, N - n_i + 1)$ is the density function of Beta distribution [1]. Since $n_i$ and $N$ are constants for bin $B_i$,

$$\int_0^{\frac{G}{S}} L(c_i)\, dc_i = \frac{1}{N+1}$$

We then normalize the likelihood function $p(n_i|c_i)$ by $\frac{1}{N+1}$ so that the sum of the normalized likelihood function becomes 1. We also replace $c_i$ with $p_i = \frac{c_iS}{G}$, probability of a read aligned to a bin with ploidy factor $c_i$. The normalized the likelihood function is denoted as $q(n_i|p_i)$, i.e.,

$$q(n_i|p_i) = f(p_i, n_i + 1, N - n_i + 1), 0 \le p_i \le 1$$

We define $1 - \alpha$ as the cumulative normalized likelihood $q(n_i|p_i)$ between lower boundary $p_{min}$ and upper boundary $p_{max}$. i.e.,

$$\int_0^{p_{\min}} q(n_i|p_i)\, dp_i = \frac{\alpha}{2}$$

and

$$\int_{p_{\max}}^{1} q(n_i|p_i)\, dp_i = \frac{\alpha}{2}$$

The boundaries can be obtained with quantiles from the beta distribution [2]:

$$p_{\min} = B\left(\frac{\alpha}{2}; n_i, N - n_i + 1\right)$$

and

$$p_{\max} = B\left(1 - \frac{\alpha}{2}; n_i, N - n_i + 1\right)$$

Then the lower and upper bound of $c$ can be obtained through $p_{\min}$ and $p_{\max}$, i.e.,

$$c^{\min} = p_{min}\frac{G}{S}$$

$$c^{\max} = p_{max}\frac{G}{S}$$

The relationship between the number of sequenced reads in a bin and its range are illustrated in Fig. S3c.

#### 1.3. Merging bins into segments

Since CNV can influence regions up to a whole chromosome, adjacent bins with similar CN values can be merged into the same segment. We assume that alignment processes to different bins are independent from each other. We want to obtain the probability that their ploidy factor difference is smaller than $\Delta$.

Assume that $n_i$ and $n_{i+1}$ reads are respectively observed to be aligned to adjacent bins $B_i$ and $B_{i+1}$ of the same size. The bivariate likelihood that we observe such data under conditions $c_i\left(0 \le c_i \le \frac{G}{S}\right)$ and $c_{i+1}\left(0 \le c_{i+1} \le \frac{G}{S}\right)$, the ploidy factors of $B_i$ and $B_{i+1}$ is given as the product of two independent binomials, i.e.,

$$L(c_i, c_{i+1}) = p(n_i|c_i)p(n_{i+1}|c_{i+1}) =$$

$$C(N, n_i)\left(\frac{c_i S}{G}\right)^{n_i}\left(1 - \frac{c_i S}{G}\right)^{N-n_i} C(N, n_{i+1})\left(\frac{c_{i+1} S}{G}\right)^{n_{i+1}}\left(1 - \frac{c_{i+1} S}{G}\right)^{N-n_{i+1}}$$

Please pay attention, the bivariate likelihood here is not normalized. Then the cumulative bivariate likelihood that bins $B_i$ and $B_{i+1}$ have ploidy factors smaller than $\Delta$ can be given as (Fig. S3d-f),

$$L(B_i = B_{i+1}) = \int_{c_i=0}^{G/S}\int_{c_{i+1}-c_t \ge -\Delta}^{c_{i+1}-c_t \le \Delta} L(c_i, c_{i+1})\, dc_i dc_{i+1}$$

Similarly, the cumulative bivariate likelihoods that the CNV of $B_i$ smaller and larger than $B_{i+1}$ can be given as (Fig. S3d-f),

$$L(B_i < B_{i+1}) = \int_{c_i=0}^{G/S}\int_{c_{i+1}-c_t \ge \Delta}^{c_{i+1}-c_t \le G/S} L(c_i, c_{i+1})\, dc_i dc_{i+1}$$

$$L(B_i > B_{i+1}) = \int_{c_i=0}^{G/S}\int_{c_{i+1}-c_t \ge -G/S}^{c_{i+1}-c_t \le -\Delta} L(c_i, c_{i+1})\, dc_i dc_{i+1}$$

The segmentation decision can be made based on the likelihood values of these three conditions, i.e., $L(B_i = B_{i+1})$, $L(B_i < B_{i+1})$, and $L(B_i > B_{i+1})$.

After the initial segmentation, the breakpoints are biased by the initial bin setting. We then generate the two bins next to each breakpoint into 5 sub-bins and optimize the breakpoints based on the read counts of these sub-bins using the same method described above. We used $\Delta$ of 0.15 in this paper.

#### 1.4. Resolution of CNVs

Assuming that $n_i$ and $n_{i+1}$ reads are respectively aligned to two adjacent bins $B_i$ and $B_{i+1}$, where $B_i$ is normal and $B_{i+1}$ contains amplification/deletion smaller than the bin size. Such CNV can be detected if bin $B_i$ can be distinguished from $B_{i+1}$. We ran a simulation with 150,000 reads (a typical read number within 1 hour sequencing using MinION), a bin size of 1Mbps (Fig. S3 g-h).

#### 1.5. Range of segmented CNVs

In Section 1.2, we obtained the CNV range for each bin. We then calculate the range of CNVs for each segment using Bonferroni correction to adjust for multiple bins. Let's assume that $m$ bins are merged into one segment. We calculate the lower and upper boundaries of CN so that cumulative normalized likelihood between the boundaries is $1-\frac{\alpha}{m}$, for each bin $B_i$, $i \in [1, m]$, based on the description in Section 1.2. We respectively denote the lower and upper boundaries of each bin's CNV as $c_{B_i}^{\min}$ and $c_{B_i}^{\max}$. Then the lower and upper boundaries of this segment is respectively given by the $c_{Seg}^{\min} = min\left(c_{B_1}^{\min} \dots c_{B_k}^{\max}\right)$ and $c_{Seg}^{\max} = max\left(c_{B_1}^{\max} \dots c_{B_k}^{\max}\right)$. Then the probability that the CNVs of all the bins within this segment is outside the boundaries is given by:

$$P\left(c_{Seg} \notin \left[c_S^{\min}, c_S^{\max}\right]\right) = 1 - \prod_{i=1}^{m} P\left(c_i \in \left[c_S^{\min}, c_S^{\max}\right]\right) \leq 1 - \prod_{i=1}^{m} P\left(c_i \in \left[c_{B_i}^{\min}, c_{B_i}^{\max}\right]\right) = 1 - \left(1-\frac{\alpha}{m}\right)^m \approx \alpha$$

#### 1.6. Likelihood of co-amplification/co-deletion

We assume that the co-amplification (co-deletion) of two chromosome arms are of interest and that the log2 ratio CNV larger (smaller) than 0.3 (-0.3) can be considered as amplification (deletion). We assume that a chromosome arm $chr_m$ contains $m$ bins denoted by $B_i$, $i \in [1, m]$ and $chr_r$ is a subset of $chr_m$ containing $r$ out of $m$ bins in the chromosome arm.
For any bin $B_i$, the cumulative normalized likelihood of amplification for $B_i$ should be,

$$L\left(CNV_{B_i} > 0.3\right) = \int_{p_{0.3}}^{1} q(p_i|t)\, dt$$

where $p_{0.3}$ is the likelihood of a random read originating from a bin $B_i$ with a log2 ratio CN of 0.3. And the the cumulative normalized likelihood of deletion for $B_i$ should be,

$$L\left(CNV_{B_i} < -0.3\right) = \int_{0}^{p_{-0.3}} q(p_i|t)\, dt$$

where $p_{-0.3}$ is the likelihood of a random read originating from a bin with a log2 ratio CN of -0.3. We assume that a chromosome arm is amplified or deleted if more than $\beta (0 \leq \beta \leq 1)$ of the arm is amplified. Then the likelihood that the chromosome arm is amplified or deleted in given by,

$$L^{AMP}_{chr_m} = \sum_{r \geq \beta m} \prod_{chr_r \subseteq chr_m} L(CNV_{B_i} > 0.3) L\left(CNV_{B_j} < 0.3\right), B_i \in chr_r, B_j \notin chr_r$$

and

$$L^{DEL}_{chr_m} = \sum_{r \geq \beta m} \prod_{chr_r \subseteq chr_m} L(CNV_{B_i} < -0.3) L\left(CNV_{B_j} > -0.3\right), B_i \in chr_r\, , B_j \notin chr_r$$

We assume that another chromosome arm $chr_n$ contains $n$ bins denoted by $B_j$, $j \in [1, n]$. Then co-amplification or co-deletion probability of these two chromosome arms are respectively given by $L^{AMP}_{chr_m} L^{AMP}_{chr_n}$ and $L^{DEL}_{chr_m} L^{DEL}_{chr_n}$.

#### 1.7. Probability of <u>no</u> co-amplification/co-deletion

Let's assume that **<u>no</u>** amplification (**<u>no</u>** deletion) of a chromosome arm is of interest. Similarly, we assume that the chromosome arm $chr_m$ contains $m$ bins denoted by $B_i$, $i \in [1, m]$. For any bin $B_i$, the cumulative normalized likelihood that a bin has **<u>no deletion</u>** should be

$$L(CNV_{B_i} > 0) = \int_{p_0}^{1} q(p_i|t)\, dt$$

where $p_0$ is the likelihood of a random read originating from a diploid bin. And the likelihood that a bin has **<u>no amplification</u>** should be:

$$L(CNV_{B_i} < 0) = \int_{0}^{p_0} q(p_i|t)\, dt$$

Assuming that the chromosome arm $chr_m$ has **<u>no amplification</u>** (**<u>no deletion</u>**) if more than $\gamma (0 \leq \gamma \leq 1)$ of the arm has **<u>no amplification</u>** (**<u>no deletion)</u>**. Then the likelihood that a chromosome arm has **<u>no amplification</u>** or **<u>no deletion</u>** is respectively given by

$$L^{NO_{AMP}}_{chr_m} = \sum_{r \geq \gamma m} \prod_{chr_r \subseteq chr_m} L(CNV_{B_i} < 0) L(CNV_{B_i} > 0), B_i \in chr_r, B_j \notin chr_r$$

and

$$L^{NO_{DEL}}_{chr_m} = \sum_{r \geq \gamma m} \prod_{chr_r \subseteq chr_m} L(CNV_{B_i} > 0) L(CNV_{B_i} < 0), B_i \in chr_r\, , B_j \notin chr_r$$

Then the likelihood that there is **<u>no</u> co-amplification/co-deletion** of two chromosomes $chr_m$ and $chr_n$ would be

$$1 - \left(1 - L^{NO_{AMP}}_{chr_m}\right)\left(1 - L^{NO_{AMP}}_{chr_n}\right)$$

and

$$1 - \left(1 - L^{NO_{DEL}}_{chr_m}\right)\left(1 - L^{NO_{DEL}}_{chr_n}\right)$$

#### 1.6. Likelihood of homozygous deletion

We assume that the homozygous deletion of a gene is of interest and that the log2 ratio CNV smaller than -0.7 can be considered as homozygous deletion and CNV larger than -0.3 can be considered as no homozygous deletion.

For any bin $B_i$, the cumulative normalized likelihood of homozygous deletion for $B_i$ should be,

$$L\left(CNV_{B_i} < -0.7\right) = \int_0^{p_{-0.7}} q(p_i|t)\,dt$$

where $p_{-0.7}$ is the likelihood of a random read originating from a bin $B_i$ with a log2 ratio CN of -0.7.

#### Section 2. Mappability and binning the genome

For a given bin size $S$, each chromosome arm of CHM13v2 reference genome is evenly split into $[S_{chr.arm}/S]$ bins, where $S_{chr.arm}$ is the size of a chromosome arm $chr \in \{chr1 \dots chrX\}, arm \in \{p,q\}$. We partitioned each chromosome of the CHM13v2 reference genome into $S = 5$ Mbps bins and then applied a sliding window of 500 bps, 1 kbps, 5 kbps, and 10 kbps to each chromosome to sample overlapping DNA sequences at intervals of 20 bps, 50 bps, 250 bps, and 250 bps, respectively. The sampled DNA sequences are aligned to the reference genome to determine whether they can be mapped back to their original locations. We observed that the centromeres are difficult to map regardless of the read length and that the telomeres are difficult to map using short reads (Fig.S2a). Therefore, we excluded centromeres and telomeres in this study.

Then each chromosome arm of CHM13v2 reference genome is evenly split into $\left[S'_{chr.arm}/S\right]$ bins ($S = 5$ Mbps), where $S'_{chr.arm}$ is the size of a chromosome arm excluding the telomeres and centromeres $chr \in \{chr1 \dots chrX\}, arm \in \{p,q\}$. Similarly, we applied a sliding window of 200bps, 500bps, 1kbps, 2kbps, 3kbps, 4kbps to each chromosome arm to sample overlapping DNA sequences at intervals of 5bps, 20bps, 50bps, 100 bps, 150bps and 250 bps, respectively. The sample reads are aligned back to the reference genome to avaluate the mappability. We found that the increased read length can decrease the mapping difficulty (Fig. S2b). Because the number of reads can influence the ranges of the CNV results, a trade-off can be found between read length/mappability and the number of sequenced reads/CNV ranges. Short reads can also decrease the total yield. An ideal read length can be designed for optimal outcomes of intraoperative CNV characterization. Our mappability analysis demonstrated that the mappability issue can be neglected with read length above 2kbps when using the CHM13v2 reference genome (Fig. S2).

We then respectively partitioned each chromosome arm of the CHM13v2 reference genome (telomeres and centromere excluded) into bins of 3Mbps for CNV characterization in this paper to balance resolution and accuracy.

**Section 3. Coverage comparison between CNV characterization using NGS and nanopore sequencing**

The yield of a typical single NGS run exceeds 30x coverage. Assuming an average read length of $150\, bps/read$, a bin of 10kbps can have a read count of:

$$\frac{30 \times 10\, kpbs/bin}{150\, bps/read} = 2k\, reads/bin$$

Long-read sequencing, an average read length of $10\, kbps/read$ would require a coverage of:

$$\frac{2k\, reads/bin \times 10\, kbps/read}{10\, kpbs/bin} = 2000x$$

to achieve the same read count per bin size.

### Section 4. Comparison of segmentation methods

Circular binary segmentation [3] was the most widely used segmentation algorithm proposed to address the problems of fluctuating CNVs values of neighboring bins in NGS or neighboring SNPs (single nucleotide polymorphisms) in arrays. It identifies CNV breakpoints through statistics:

$$Z_C = Max_{1 \le i < j \le n} \left| Z_{i,j} \right|$$

where

$$Z_{i,j} = \{1/(j-i) + 1/(n-j+i)\}^{1/2} \cdot \{\frac{S_j - S_i}{j-i} - \frac{S_n - S_j + S_i}{n-j+i}\}$$

DNAcopy was intentionally designed to incorporate a ‘penalty term’ of $j - i$ so that it rather merged neighboring regions than breaking them so as to reduce the effect from fluctuating CNV values in a small region. However, this method can cause unreliable and inaccurate CNV results when the bin size is set to be Mbps since neighboring bins with different CNVs tend to be merged.

### Reference


1. Gupta, A.K. and S. Nadarajah, *Handbook of beta distribution and its applications*. 2004: CRC press.
2. Måns, T., *The cost of using exact confidence intervals for a binomial proportion.* Electronic Journal of Statistics, 2014. **8**(1): p. 817-840.
3. Olshen, A.B., et al., *Circular binary segmentation for the analysis of array‐based DNA copy number data.* Biostatistics, 2004. **5**(4): p. 557-572.
